# Using electronic health record data accessed via OpenSAFELY to develop indicators of end-of-life care quality

**DOI:** 10.64898/2026.02.03.26345473

**Authors:** Stuti Bagri, Sophie Julian, Miranda Davies, Sarah Scobie, Andrea Schaffer, The OpenSAFELY Collaborative

## Abstract

An understated disruption to health services brought about by the Covid-19 pandemic was the increase in deaths occurring outside a hospital. Since quality of end-of-life care is typically monitored through place of death and hospital activity, a new approach focused on care in community settings is needed. In this study, we aimed to test whether patient-centric measures of quality at the end of life can be derived from primary care electronic records.

With the approval of NHS England, analysis was undertaken in OpenSAFELY-TPP using electronic health care records of over 970,000 patients who died between March 2019 and August 2023, covering periods before, during and after the pandemic. We developed two new measures of end-of-life care quality—specialist palliative care team contacts and advance care planning, and tracked the proportion of patients with these records, categorized by place and cause of death, along with an existing measure indicating palliative care needs.

The proportion of people with a GP record of specialist palliative care was 4-5% on average, higher for those who died of cancer or died in a hospice. Advance care planning records increased from 19% to 27% (barring a decrease following the onset of the Covid-19 pandemic) driven in large part by increases for patients who died in care homes. Advance care planning and recording of palliative care needs were plausible measures to track changes in care, unlike the specialist palliative care measure where recorded use was sparse. Improved coding in primary care records would improve reliability of measures.

**Key messages:**

- Quality of end-of-life care is traditionally measured by how patients use health services (for example emergency department attendances)
- We used routine GP health records to track aspects of end-of-life care quality which matter to patients and discuss the impact of the covid-19 pandemic on these quality measures
- A new measure of advance care planning and the existing palliative care needs measure could be used to track end of life care delivered in the community
- The measure of specialist palliative care was sparsely coded and unlikely to be useful unless coding and data linkage between GP and other systems improves

## Introduction

Life expectancy in the United Kingdom has increased over the last four decades(1), while the number of people living with multiple long-term conditions has increased, with 68% of adults aged 80 or older having two or more long-term conditions(2). Collectively, this creates an increase in demand for end-of-life care services while simultaneously making end of life care more complex to deliver. While there is a large body of evidence regarding what patients value – such as sufficient pain relief – there can be a gap between patient experience and the reality(3).

The COVID-19 pandemic disrupted healthcare services globally and the place where people died changed significantly in the UK. A third more people died at home during 2020-2022 than the pre-pandemic five-year average and the proportion of home deaths has since remained above pre-pandemic levels(4).

We established a project to evaluate the changes in care at the end of life and describe trends in where people were dying. We found significant changes in care for people who died at home (4), in care homes(5), and in the use of hospital services(6). This work was in line with WHO recommendation for COVID-19 to rapidly assess health care delivery and develop key performance indicators to quantitatively evaluate care (7).

We used previously published indicators for our initial work(8), however due to the substantial changes in clinical care, and rapid deployment of technology(9) we identified a need to assess whether new indicators could be developed to support monitoring the ongoing impact of the COVID-19 pandemic in line with developments in GP data.

Changes in place of death during the pandemic highlighted gaps in existing routine data on end of life of care, which has been focused on hospital care and measures of activity, for example emergency admissions at the end of life(10). Patient centred outcomes are not yet routinely used when evaluating the quality of end-of-life care(11). With the reduction in the proportion of deaths occurring in hospital persisting post pandemic, routine monitoring of end-of-life care quality in primary care is essential to ensure that patients are receiving the care they need, and to understand the implications for primary and community service capacity.

In this paper, we describe current indicators of measuring care at the end of life, investigate patterns of recording care at the end of life in GP electronic records, and propose and assess new measures of care using linked GP, hospital and death registration data. We consider the influence of COVID-19 on the trends identified by these new measures.

## Methods

### Developing measures

To develop measures of quality of care we reviewed literature to identify dimensions of good quality end-of-life care.

These included effective symptom management, such as timely access to medication and effective pain relief (12), coordination of services, and good communication between patients and their family members and healthcare (13). A lack of integration between services is a barrier to good quality care(14).

Receiving care from health care staff with specific expertise in palliative and end of life care is valued by patients and families, as specialist palliative care team contacts can have an important role in symptom control, especially around pain management(15). Continuity of contact with the specialist palliative care team is also vital in order to build trusted relationships(16).

At its core, good quality end of life care requires consulting with patients on what matters to them(17), and giving them autonomy in choosing how they are being looked after, for which a precondition is recognition that someone is approaching the end of life (11). Advance care planning refers to the process by which patients can state preferences about their treatment enabling their wishes to be respected if they lose decision making capacity.

Based on these themes, we identified six potential measures of good quality end of life care: a) recognition of end-of-life b) advance care planning c) continuity of care d) co-ordinated services e) specialist palliative care team contacts f) anticipatory medicine. To determine the feasibility of creating measures, we curated a preliminary list of corresponding SNOMED codes which are used by GPs in patients’ electronic health records, using data published by NHS Digital (18) to identify key codes relating to each measure and recorded use of these codes between August 2021 to July 2022. Using this information, we consulted with our advisory group on which measures could be feasibly developed.

For measures considered feasible, we then curated a comprehensive codelist, including all SNOMED codes we deemed relevant to each measure. Codelists were developed using OpenCodelists, a tool which allows researchers to build their own set of SNOMED codes for use on OpenSAFELY projects. The full code lists can be found as a supplemental file in the appendix.

OpenSAFELY is a platform which enables analysis of GP electronic health records in England. These records are digital logs of interactions and actions related to patients in a primary care setting. They can include things like medication prescriptions and clinical decisions which are coded (using SNOMED clinical terms). However, since end of life care is usually delivered by providers in different settings, like hospitals, hospices etc, GP records may not be a complete record of all end of life care received, and information which is free text is not accessible.

### Data

For the three measures examined in detail, we used the OpenSAFELY data platform(19) to explore the health service use of patients registered with a TPP practice who died between March 2019 and August 2023. Primary care records managed by the GP software provider, TPP were linked to hospital activity and ONS death data through OpenSAFELY. March 2019 was selected as the start of the period based on when information regarding date of death was made available by the Office for National Statistics (ONS) within OpenSAFELY as part of our earlier study. Data management was performed using Python, with analysis carried out using R.

All data were linked, stored, and analysed securely using the OpenSAFELY platform, https://www.opensafely.org/, as part of the NHS England OpenSAFELY COVID-19 service. Data included pseudonymised data such as coded diagnoses, medications, and physiological parameters. No free text data are included. All code is shared openly for review and re-use under MIT open license. Detailed pseudonymised patient data is potentially re-identifiable and therefore not shared. More information is available as a supplemental file in the appendix.

### Study Population

Participants included patients registered with a TPP practice in England who died. Patients whose death was not registered by ONS and had invalid/missing values for sex were excluded. The TPP system covers the records of approximately 24 million patients across England. The dataset includes 975,125 people who died, which equates to 40% of all ONS-published deaths in England across the study’s period(20).

### Data analysis

We tracked a) the proportion of patients who died with palliative care recorded in their GP records, b) the proportion of patients who died with specialist palliative care recorded in their GP records, c) the proportion of patients who died with an advance care plan recorded in their GP records, in the 90-days prior to date of death.

Although end of life care can be provided over a longer period, 90-days was deemed a sufficient amount of time to capture majority of end of life care activity, and is also used by the Office of Health Improvement and Disparities (OHID) to report on emergency admissions in the end of life period.

Measures were further sub-categorised by place and cause of death. Place of death records if patients die at ‘Home’, in a ‘Hospice’, ‘Hospital’, ‘Care home’ or ‘Elsewhere/other’. Cause of death was categorized as ‘Cancer’, ‘Circulatory diseases’, ‘Dementia and Alzheimer’s disease’, ‘Flu and pneumonia’, ‘Other respiratory diseases’ or ‘All other causes’. Developing and tracking these measures in this way allowed us to establish their validity by comparing them to what we would expect to observe in practice.

In compliance with disclosure control policy mandated by OpenSAFELY, counts below 8 were redacted and all other numbers were rounded to the nearest five.

### Patient and public involvement

The research was supported by an advisory group including general practitioners and academics specialising in palliative care, patients in receipt of end-of-life care and family members providing support. The advisory group provided feedback on the project plan, the quality-of-care measures selected and interpreting interim results.

## Results

The reasoning behind our selection of measures is described in Table 1.

**Table 1:**
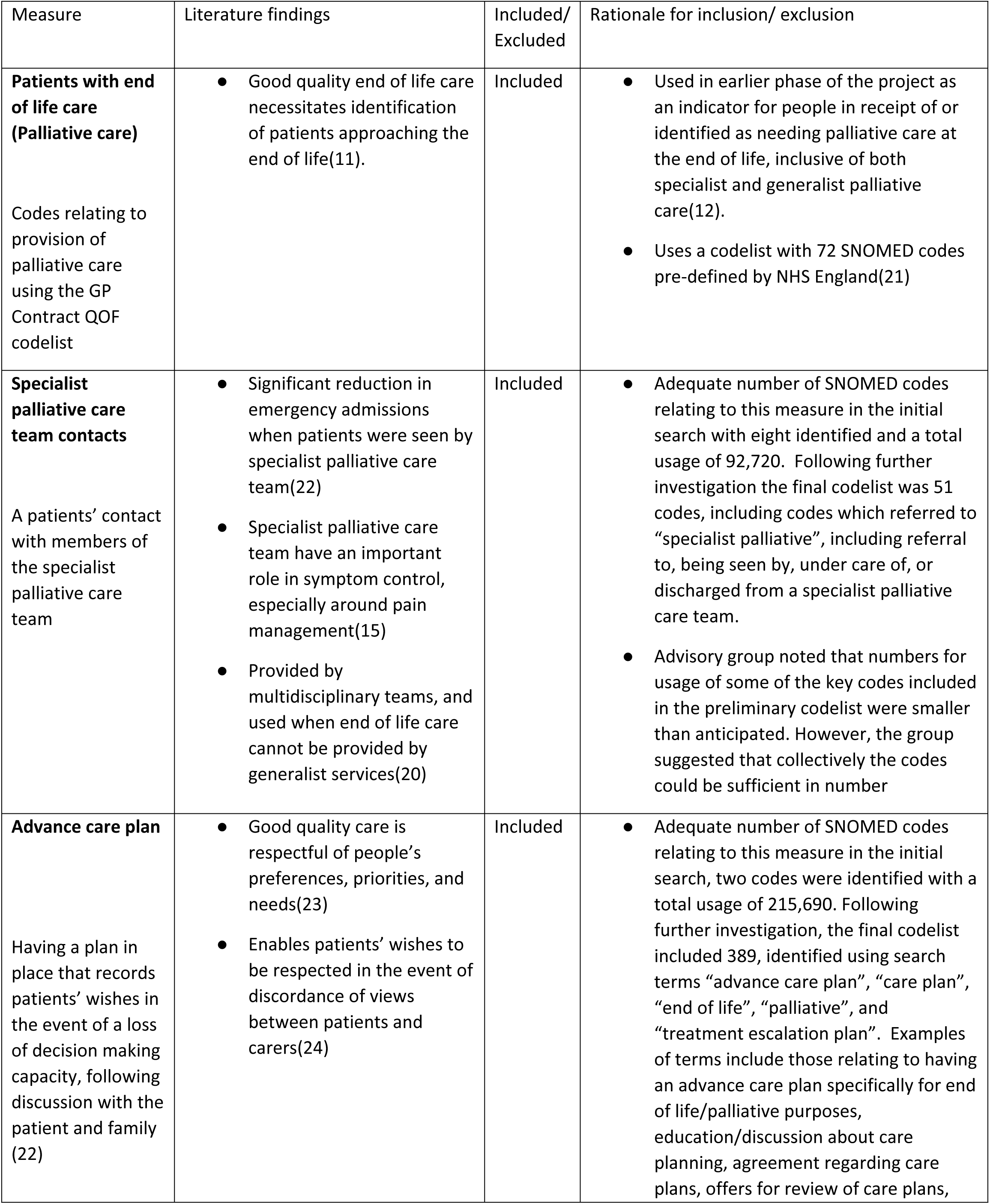
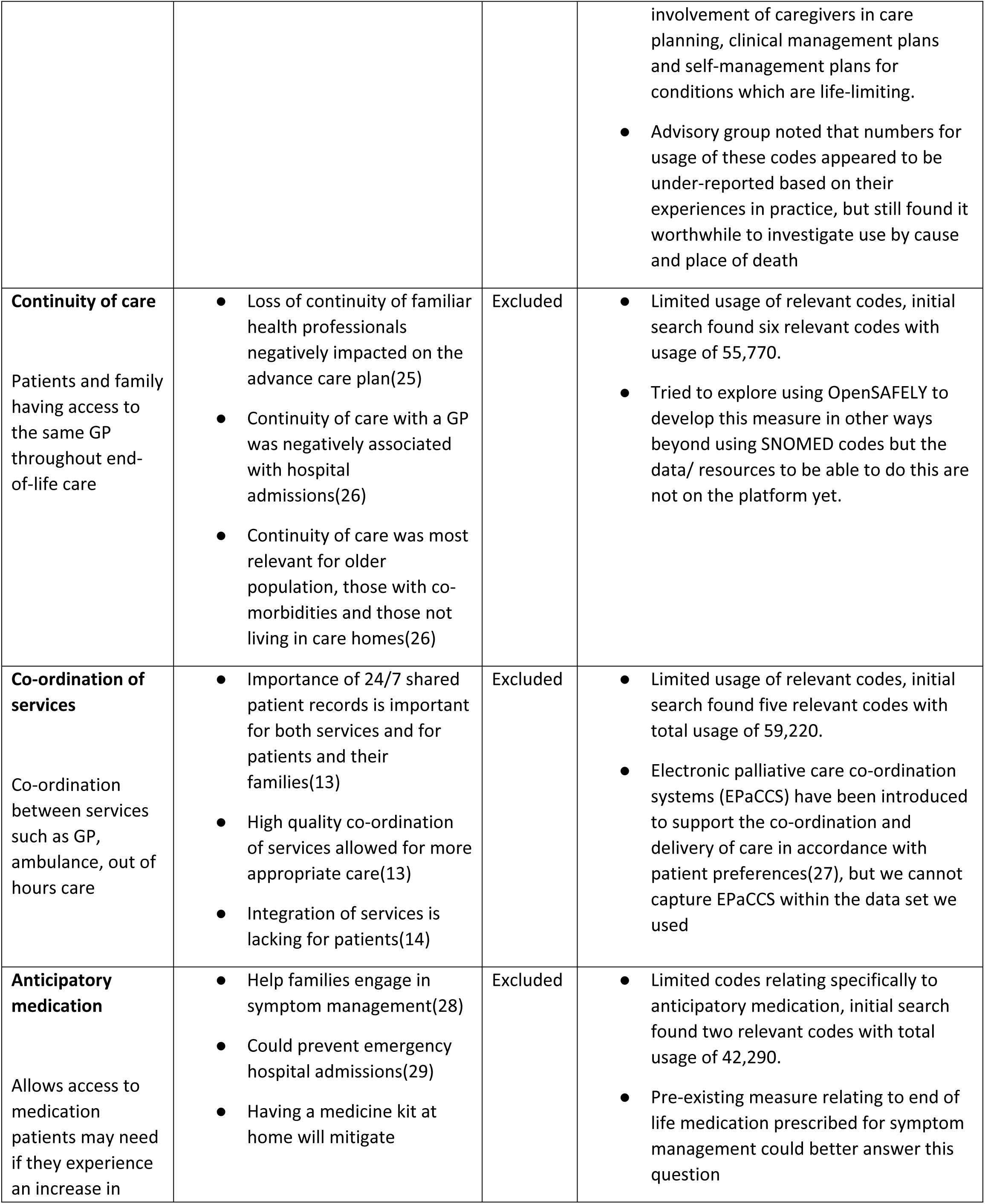
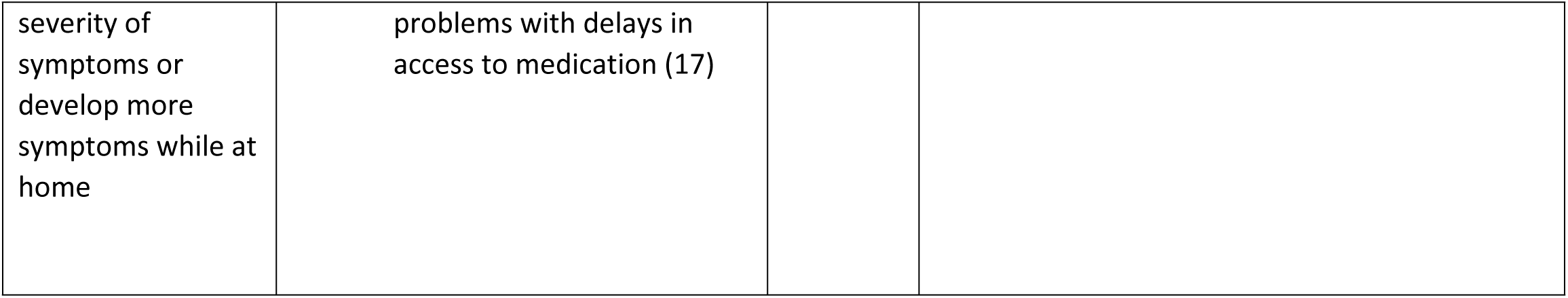
Measures considered and reasons for inclusion/exclusion.

### Place and cause of death

Deaths most frequently occurred in hospitals (43.4%), followed by homes (27.7%), care homes (21.7%), hospices (4.7%), and elsewhere/other (2.5%). Cancer was listed as the main cause of death for 25.6% of all deaths that occurred, followed by circulatory diseases (23.8%), “other causes” (21.6%), dementia and Alzheimer’s (11.4%), respiratory diseases (7.4%), Covid-19 (6.4%), and lastly, flu and pneumonia (3.5%). The profile of place and cause of death is similar to all deaths for England(20).

### Patients with end of life care (palliative care)

The proportion of people who died with palliative care recorded in their GP record rose from 23.1% to 27.7% across the timeframe. However, during the pandemic the proportion of people with palliative care recorded in their GP records decreased from 29.5% in June 2020 to 19.6% in January 2021. For patients who died in a care home there was a sharp drop in the proportion of patients who had palliative care recorded at the start of the Covid-19 pandemic, decreasing from 49.9% in February 2020 to 39.7% in April 2020 (Figure 1). Similarly, during the second wave of the Covid-19 pandemic the proportion of patients who had palliative care recorded decreased, with the most notable decrease seen in those who died in hospices. Since the pandemic, the proportion of patients who died at home who had palliative care recorded in their records has been increasing, this increase has been sustained since the end of 2022.

**Figure 1:**
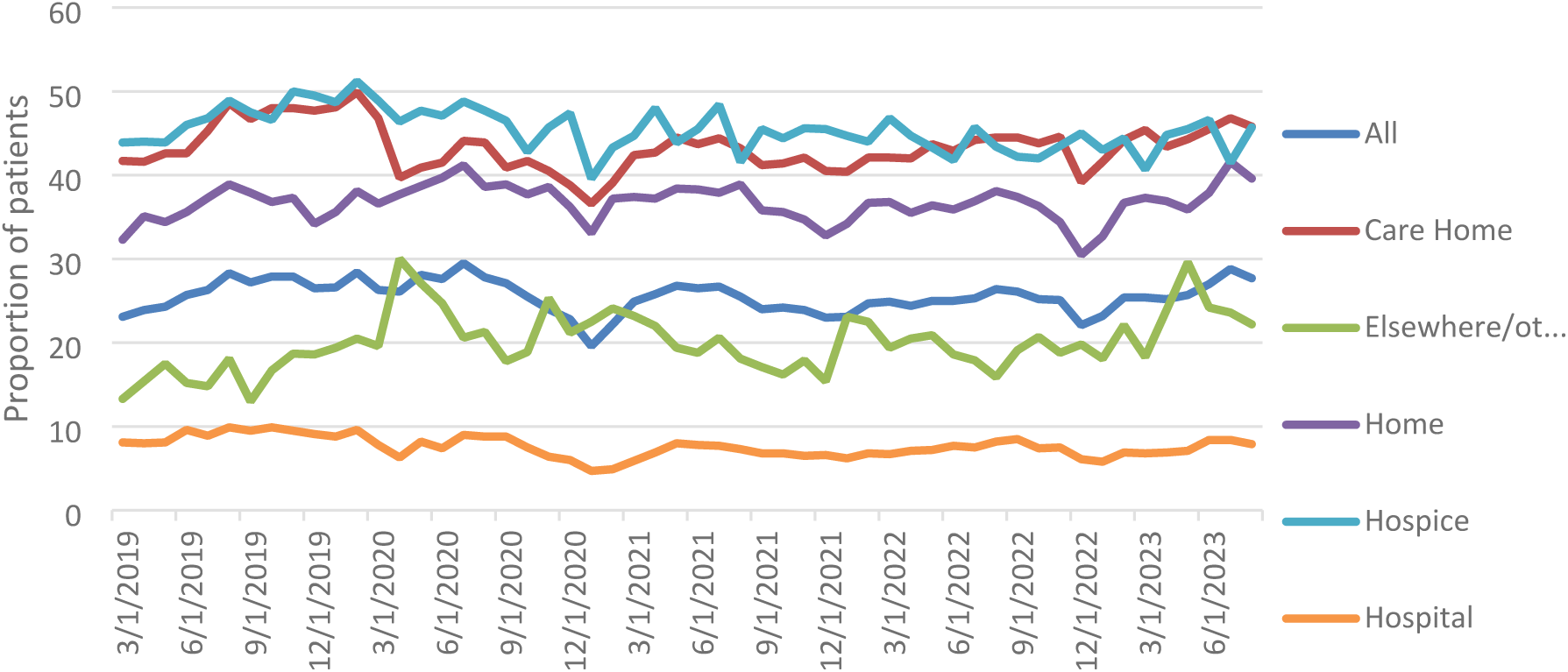
Proportion of patients who died with palliative care recorded in their GP records in the last three months of life, by place of death; March 2019–August 2023

Patients whose underlying cause of death was cancer had the largest proportion of palliative care recorded in their GP records at 45.3% in August 2023 followed by patients with dementia (38.9% in August 2023) (Figure 2). For patients who died due to other conditions, the proportion with palliative care records remained below 20% for most months.

**Figure 2:**
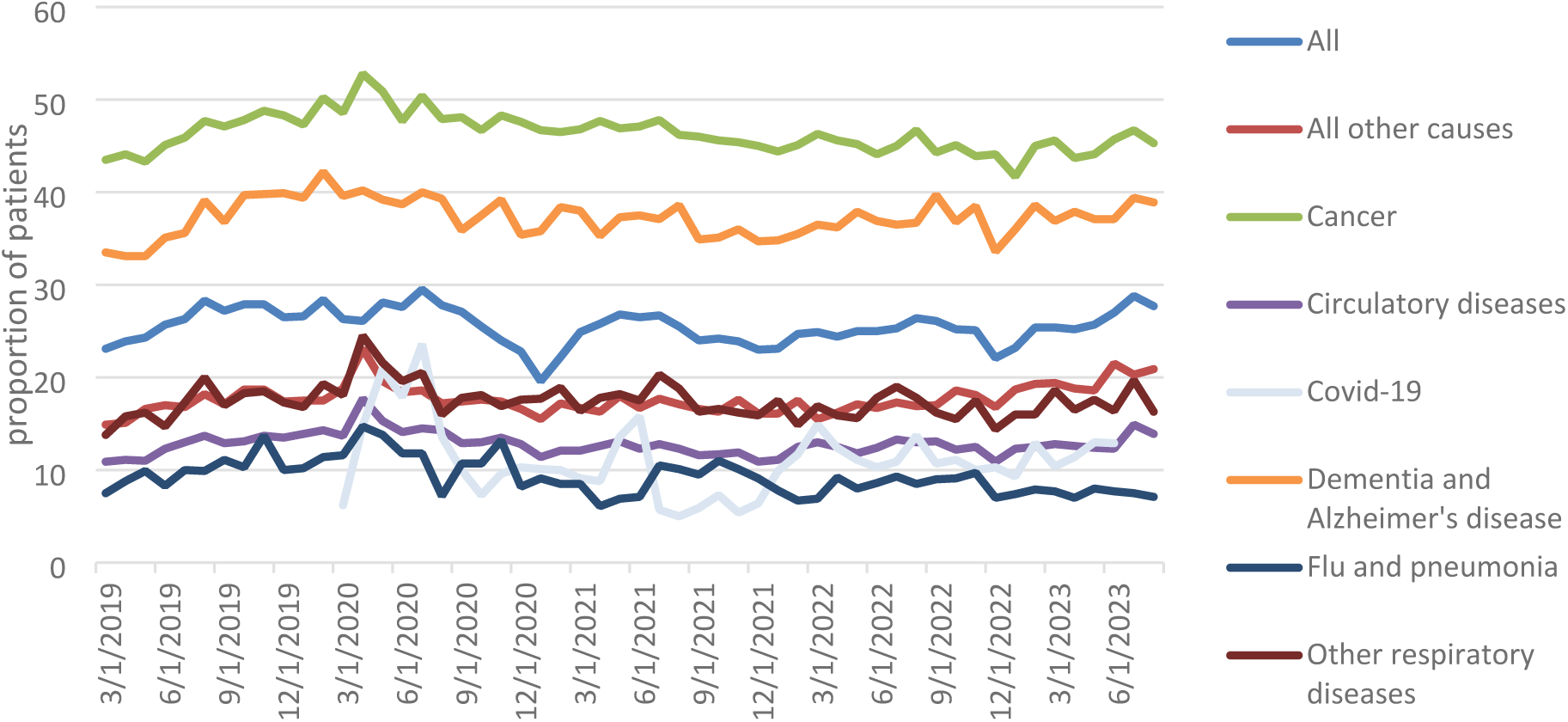
Proportion on patients who died with a record of palliative care in their GP records in the last three months of life, by cause of death; March 2019–August 2023

### Specialist palliative care

The proportion of all people who died with specialist palliative care recorded in their GP records was relatively low across the period, ranging from between 4% and 5% between March 2019 and August 2023. During the early months of the pandemic there was a slight increase in the number of people who had specialist palliative care recorded in their GP records, from 3.4% in April 2020 to 5.1% in July 2020.

Patients who died in a hospice consistently had the largest proportion of specialist palliative care contacts in their GP records, reaching 14.9% in August 2023 (Figure 3). They were followed by patients who died at home, where the proportion of specialist palliative care contacts ranged from 7.4% to 8.8% across the study period.

**Figure 3:**
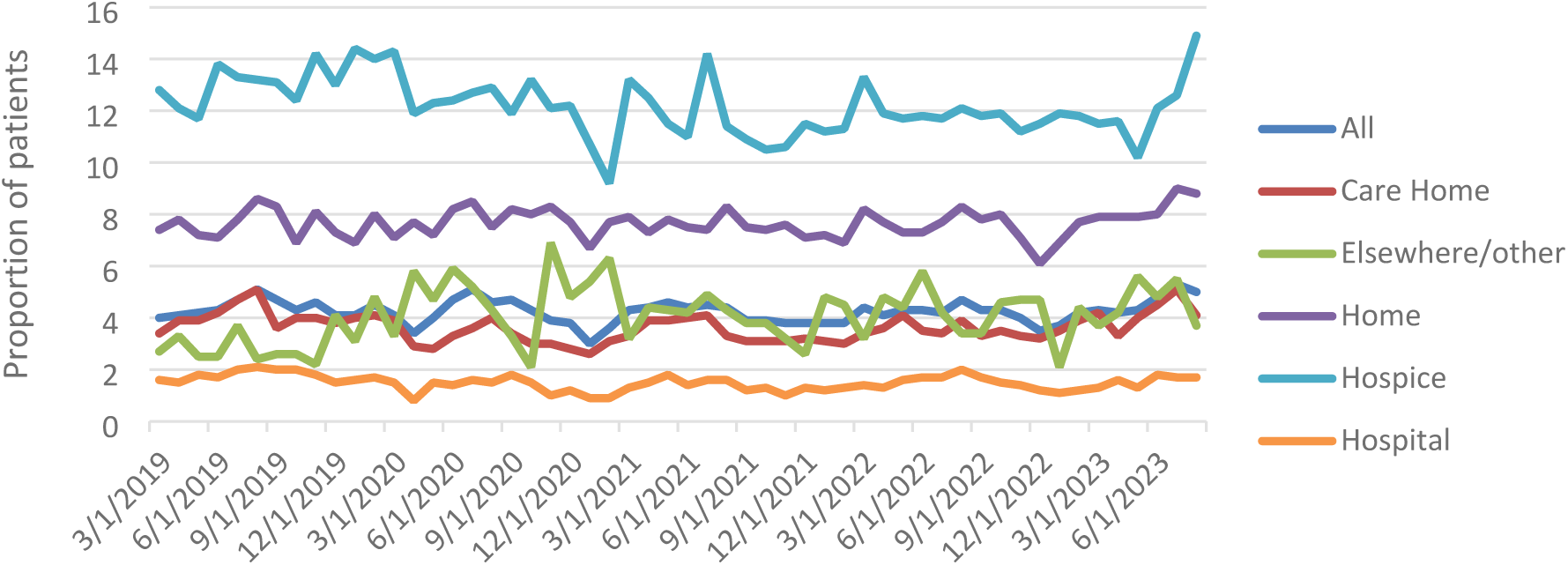
Proportion of patients who died with a record of specialist palliative care team contacts in their GP record in the last three months of life, by place of death; March 2019–August 2023

People with an underlying cause of death of cancer had the largest proportion of specialist palliative care recorded in their GP records, remaining at around 10% across the timeframe (Figure 4). For all other causes of death, this mostly remained below 5% of patients. Flu and COVID-19 are not shown due to small numbers of deaths requiring disclosivity redaction.

**Figure 4:**
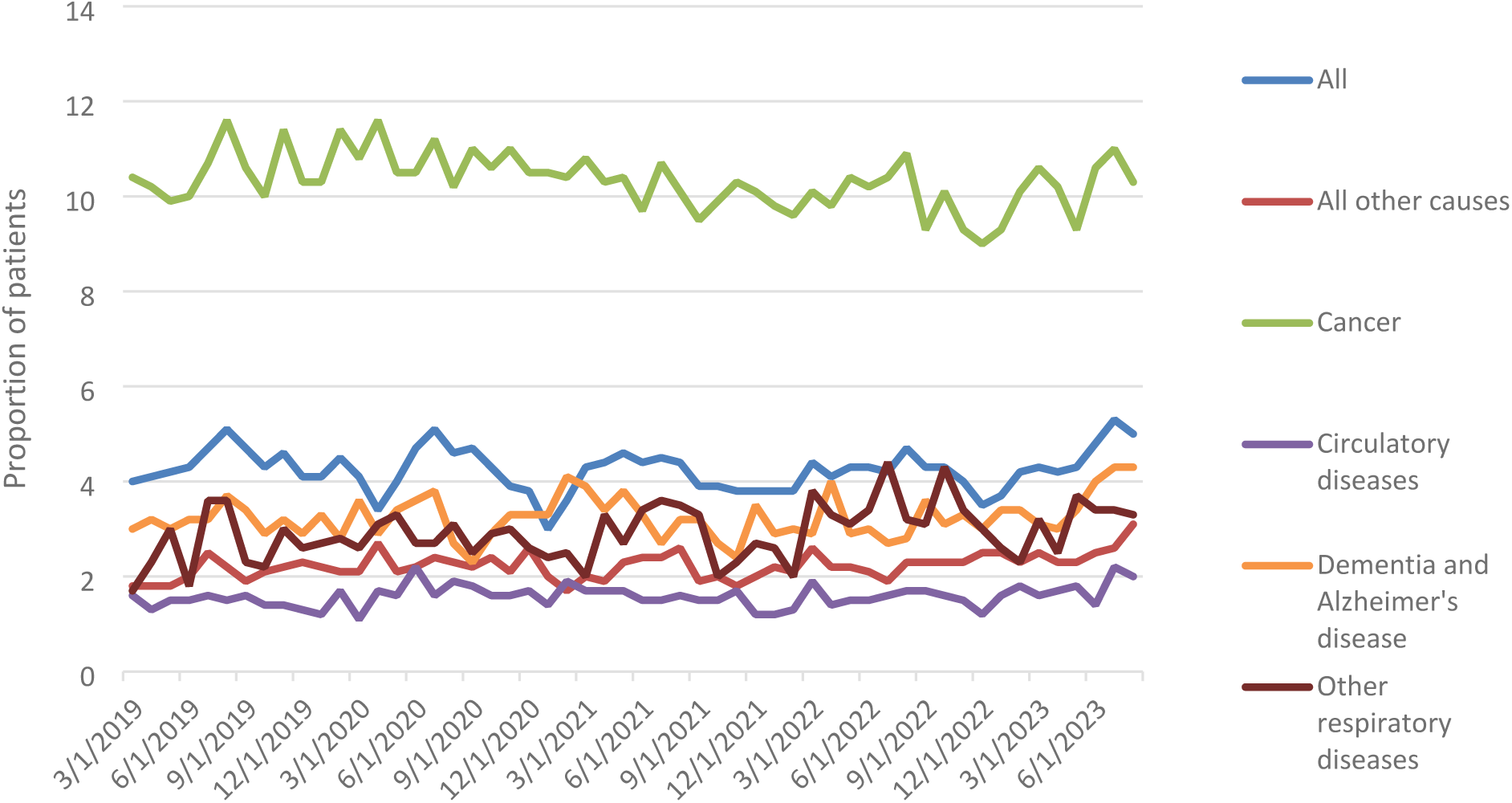
Proportion of patients who died with specialist palliative care team contacts recorded in their GP records in the last three months of life, by cause of death: March 2019–August 2023

### Advance care planning

The proportion of all people who died with advanced care planning recorded in their GP records has increased over time from 19% in March 2019, to 26.7% in August 2023, but this was driven largely by recording specific to patients who died in care homes (Figure 5). Following the onset of the COVID-19 pandemic in April 2020, advanced care planning records decreased before picking up again in early 2021. In terms of variation over time by place of death, people who died in a care home were consistently most likely to have a GP record of advance care planning. Increases over time were far less pronounced for other groups; with people who died in hospital tending to be least likely to have advance care planning recorded in their GP record. In August 2023, 13.3% of people who died in hospital had a GP record of an advanced care plan, compared to 51.1% of those who died in care homes, and 26.7% of those who died at home.

**Figure 5:**
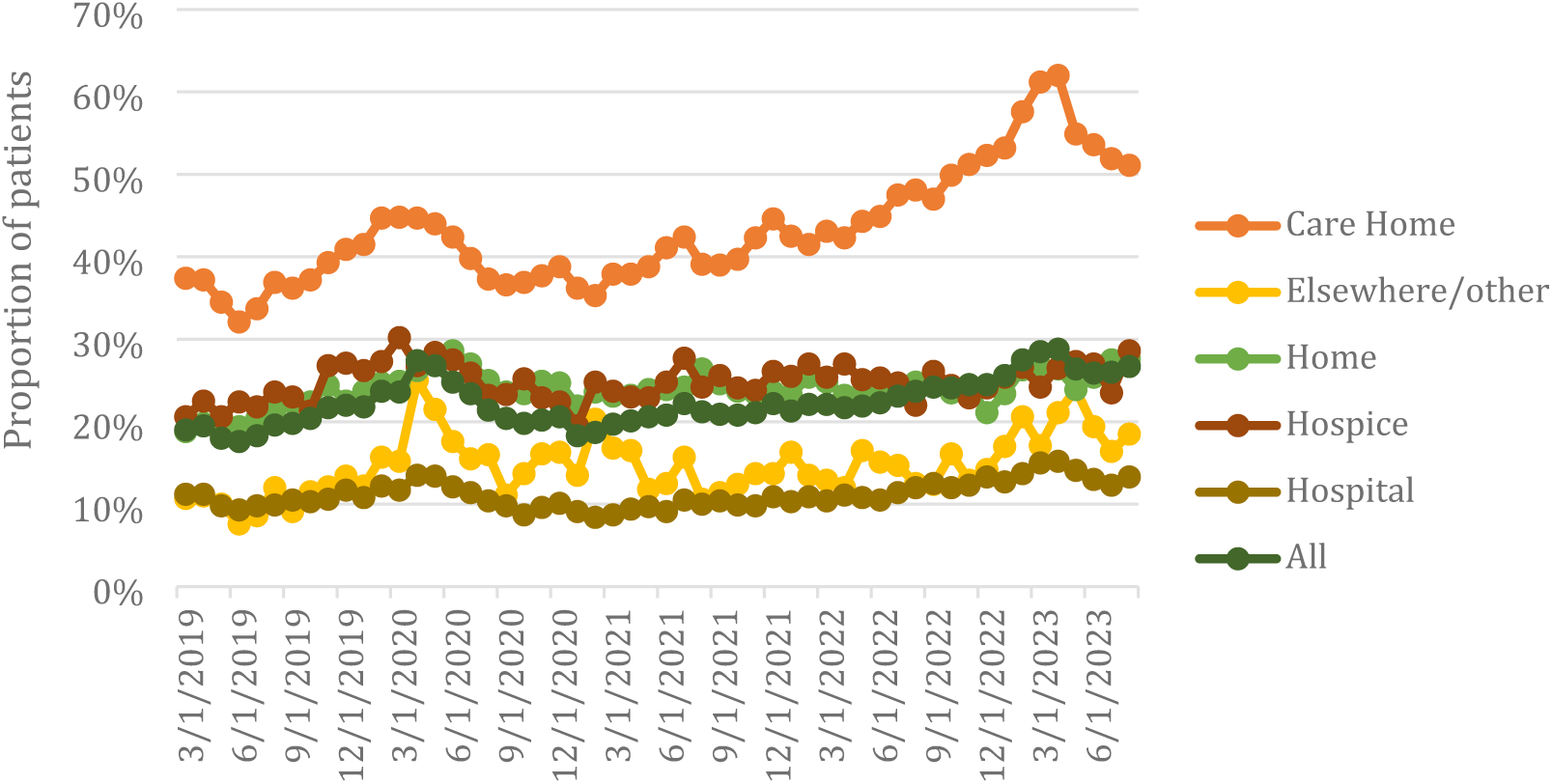
Proportion of patients who died with a record of an advance care plan in their GP record in the last three months of life, by place of death; March 2019–August 2023

People with a diagnosis of dementia or Alzheimer’s disease were most likely to have a GP record of advance care planning at any point across the period, followed by those with a cancer diagnosis (Figure 6). In April 2023, 59.4% of patients who died with a diagnosis of dementia or Alzheimer’s had a GP record of an advanced care plan, which was the highest proportion recorded across the period by any cause of death. In August 2023, 47.4% of those who died with a diagnosis of dementia or Alzheimer’s had an advanced care plan recorded on their GP record, compared to 28.8% of patients with a cancer diagnosis, and just 13.9% of patients who died of Covid-19.

**Figure 6:**
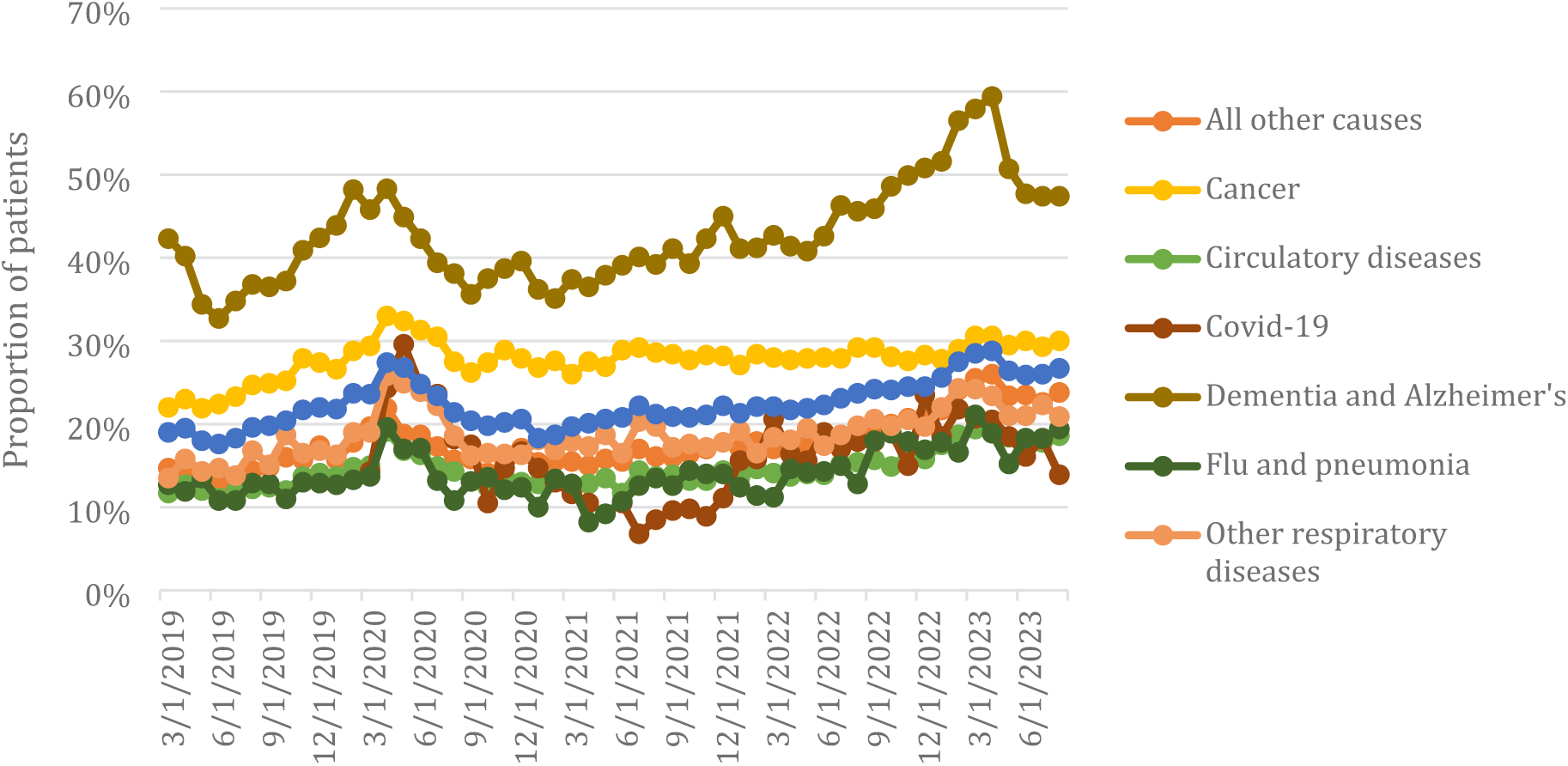
Proportion of patients who died with a record of an advanced care plan in their GP record int he last three months of life, by cause of death; March 2019–August 2023

## Discussion

This study examined the scope to use GP electronic records to develop measures of quality of end of life care, as part of a larger project to track changes in care resulting from the Covid-19 pandemic. The pandemic highlighted the need for measures of quality relating to care outside hospitals, with more people dying at home and a shift towards care in the community.

### Palliative care and specialist palliative care measures

The Covid-19 pandemic influenced the proportion of patients with palliative care recorded in their GP records. Namely, the recording of palliative care decreased following the first wave of the pandemic before levelling out and decreasing again during the second wave of the Covid-19 pandemic. Whereas there was minimal effect on specialist palliative care, likely influenced by the small proportions in this measure. Across all causes and places of death, the proportion of all patients with GP-recorded specialist palliative care contacts was consistently low, under 5% for most months. It is likely that this reflects clinical coding practice in GP records rather than the delivery of care. Feedback from the advisory group stated that most GPs group all palliative care, both generalist and specialist together using only palliative care codes in practice. This is in line with the results from the palliative care measure, where recorded use was much higher. This suggests that the previously developed palliative care measure provides a better representation of palliative care received in practice.

The underestimation of specialised palliative care services will be more pronounced in certain settings where primary care interaction is low. Patients who die in hospitals may be receiving inpatient specialist palliative care which would be recorded in their hospital record but likely not be reflected in their GP record. Notably, hospital deaths included a large proportion of Covid-19 deaths but recording of specialist palliative care codes were too small to report.

Our findings highlight some of what is already known about specialist palliative care services. Specialist palliative care is often provided locally by hospice and hospice-at-home services, and this is reflected in our findings which show that the proportion of patients who died in hospices and at home that have specialist palliative care recorded is higher than in other settings. Additionally, the long-standing pattern of end-of-life care services being predominantly focused on providing care for patients with cancer(30) was visible in the data, with patients who died of cancer most likely to receive specialist palliative care services compared to other causes of death.

### Advance care planning

The overall increase in the proportion of patients with a GP-recorded advanced care plan was in large part driven by an increase in the proportion of patients who died in care homes with a record of an advanced care plan up until April 2023. This period coincided with the enhanced health in care homes program (EHCH)(31) set up to integrate care between primary care networks and care homes, which incentivised recording personalised care plans up until April 2023. The trajectory of the proportion of patients in a care home with a GP-recorded advanced care plan matches that of patients whose main cause of death was dementia and Alzheimer’s. These patients were more likely than those who died of other clinical conditions to have a record of advance care planning, possibly because they were dying in care homes where at least a third of the residents suffer from dementia(32). There was an overall decrease in advance care planning records following the onset of the Covid-19 pandemic in March 2020 up until the end of the year, potentially because care provision was disrupted during that time. Since the end of 2020, the proportion of patients with advance care planning records has been increasing.

Although the usefulness of the measure will be hampered by its reliance on record sharing between GP practices and other services, the proportion of patients with an advance care plan recorded is plausible. Small increases in the use of advance care planning over time in all settings could represent better integration of care into practice. Those who died of acute respiratory conditions (including flu, pneumonia) may have been less likely to have an advance care plan record due to a shorter illness before death.

## Strengths and limitations

Our study developed two novel indicators of care quality at end of life which build on evidence of aspects to care which matter to patients, in keeping with the drive towards patient-centred care(33), and opportunities to use routine electronic health records to understand quality of care. The trends in use of these measures, as well as an existing measure of recognition of palliative care provide new insight into quality of care in community settings at the end of life at a time of significant change in health services and patterns of care at the end of life(7–9). The results are drawn from a large scale, nationally representative dataset covering 40% of deaths in England, making the scale of analysis unprecedented. The study also uses NHS standard coding – SNOMED – and the analysis code is openly available for re-use. The advisory group included patients in receipt of end-of-life care as well as family carers, general practitioners, academic and policy experts, who contributed into the design of the study and interpretation of the data.

The indicators do not reveal the day-to-day experience of care. For instance, while the presence of an advanced care plan is positive, the plan needs to be revised and shared appropriately between services, and followed, to be meaningful. As such, these measures should be treated as distinct from patient-reported outcome measures. Being reliant on GP data limited our access to detailed information and documentation of end-of-life processes. The Electronic Palliative Care Co-ordination Systems (EPaCCS)(34), a dataset designed for documenting palliative care activity at a regional level, may make a better resource for understanding palliative care activity with a more granular lens, if this data was incorporated into linked datasets.

### Policy implications

We identified changes in coding relating to changes in demand for services and disruption of usual services at the start of the Covid-19 pandemic, most notable for advance care planning and palliative care measures. For the palliative care measures, we continued to see an impact on recording during the second wave of Covid-19. Since the pandemic, all measures have been gradually increasing for all deaths, with differing trends for different places and causes of death. Continuing to track changes in coding in primary care is critical at a time when there is significant change in provision of services as well as use of electronic health records.

The palliative care needs and advance care planning measures both show value in understanding trends in end of life care, have face validity, and could readily be implemented in the NHS and adapted for use in different settings. The specialist palliative care measure had low levels of coding and hence limited value for monitoring quality of care.

Better coding and linkage between GP and other clinical records would result in better estimates of these measures using this methodology. This will ultimately contribute to a wider pool of knowledge on end-of-life care services in England, especially vital in a post-pandemic world where the landscape of end-of-life care delivery has changed.

## Conclusion

This work pilots the use of electronic health records data, accessed via OpenSAFELY, to construct measures of quality of end-of-life care which are meaningful to patients and relevant to end of life care delivered in community settings. Two new measures were produced, specialist palliative care team contacts and advance care planning, and their use over time by cause and place of death was tracked. Of the two, advance care planning was deemed a more useful measure because it was more likely to accurately reflect care activity, as opposed to provider coding practices.

## Acknowledgements

We are very grateful for all the support received from the TPP Technical Operations team throughout this work, and for generous assistance from the information governance and database teams at NHS England and the NHS England Transformation Directorate. We are also thankful to our advisory group, whose input was of immense value to this study.

The OpenSAFELY Collaborative involves the following people:

Dr. Amelia Green, Amir Mehrkar, Ben Goldacre, Ben Butler-Cole, David Evans, George Hickman, Dr. Iain Dillingham, Dr. Jon Massey, Louis Fisher, Lucy Bridges, Dr. Milan Wiedemann, Peter Inglesby, Dr. Rebecca Smith, Sebastian Bacon, Dr. Simon Davy, Dr. Steven Maude, Thomas O’Dwyer, Tom Ward, Liam Hart, Pete Stokes, Dr. Christopher Bates, Jonathan Cockburn, John Parry, Frank Hester, Sam Harper.

The advisory group consisted of the following people:

Dr Sarah Mitchell (Chair). Interim National Clinical Director for End-of-Life Care, NHS England & Clinical GP in Sheffield. Clinically active general practitioner.

Professor Fliss Murtagh. Professor of Palliative Care and Director of the Wolfson Palliative Care Research Centre; Hull York Medical School, University of Hull Wolfson Palliative Care Research Centre

Brian McKenna. Honorary Research Fellow EBM Datalab, OpenSAFELY

Katie Griffin. Patient and Public Involvement representative

Ann-Marie Wilson. Patient and Public Involvement representative

Jude Beng. Patient and Public Involvement representative

Dr Karen Chumbley. Chief Clinical Officer Clinical Lead in End-of-Life Care, St Helena Hospice Suffolk and North East Essex. Clinically active general practitioner.

Jo Greengrass. Essential Standards of Care Quality Improvement Lead, NHS Frimley Health and Care ICS

## Competing Interests Statement

The OpenSAFELY platform is principally funded by grants from:

● NHS England [2023-2025];
● The Wellcome Trust (222097/Z/20/Z) [2020-2024];
● MRC (MR/V015737/1) [2020-2021].

Additional contributions to OpenSAFELY have been funded by grants from:

● MRC via the National Core Study programme, Longitudinal Health and Wellbeing strand (MC_PC_20030, MC_PC_20059) [2020-2022] and the Data and Connectivity strand (MC_PC_20029, MC_PC_20058) [2020-2022];
● NIHR and MRC via the CONVALESCENCE programme (COV-LT-0009, MC_PC_20051) [2021-2024];
● NHS England via the Primary Care Medicines Analytics Unit [2021-2024].

The views expressed are those of the authors and not necessarily those of the NIHR, NHS England, UK Health Security Agency (UKHSA), the Department of Health and Social Care, or other funders. Funders had no role in the study design, collection, analysis, and interpretation of data; in the writing of the report; and in the decision to submit the article for publication.

## Data availability statement

All data relevant to the study are included in the article or uploaded as supplementary information. All code for data management and analyses are archived at: https://github.com/opensafely/end-of-life-carequality. All the code lists we used in the analysis are publicly available on the projects [GitHub page](https://github.com/opensafely/end-of-life-carequality/tree/main/codelists). Access to the data platform is via a virtual private network connection and restricted to a small group of researchers.

## Rights retention statement

For the purpose of Open Access, the author has applied a CC BY public copyright licence to any Author Accepted Manuscript (AAM) version arising from this submission.

## Information governance and ethical approval

NHS England is the data controller of the NHS England OpenSAFELY COVID-19 Service; TPP is the data processor; all study authors using OpenSAFELY have the [approval of NHS England](https://digital.nhs.uk/coronavirus/coronavirus-covid-19-response-information-governance-hub/the-nhs-england-opensafely-covid-19-service-privacy-notice). This implementation of OpenSAFELY is hosted within the TPP environment which is accredited to the ISO 27001 information security standard and is [NHS IG Toolkit compliant](https://digital.nhs.uk/data-and-information/looking-after-information/data-security-and-information-governance/data-security-and-protection-toolkit).

Patient data has been pseudonymised for analysis and linkage using industry standard cryptographic hashing techniques; all pseudonymised datasets transmitted for linkage onto OpenSAFELY are encrypted; access to the NHS England OpenSAFELY COVID-19 service is via a virtual private network (VPN) connection; the researchers hold contracts with NHS England and only access the platform to initiate database queries and statistical models; all database activity is logged; only aggregate statistical outputs leave the platform environment following best practice for anonymisation of results such as [statistical disclosure control](https://digital.nhs.uk/data-and-information/information-standards/information-standards-and-data-collections-including-extractions/publications-and-notifications/standards-and-collections/isb1523-anonymisation-standard-for-publishing-health-and-social-care-data) for low cell counts.

The service adheres to the obligations of the UK General Data Protection Regulation (UK GDPR) and the Data Protection Act 2018. The service previously operated under notices initially issued in February 2020 by the Secretary of State under Regulation 3(4) of the Health Service (Control of Patient Information) Regulations 2002 (COPI Regulations), which required organisations to process confidential patient information for COVID-19 purposes; this set aside the requirement for [patient consent](https://www.gov.uk/government/publications/coronavirus-covid-19-notification-of-data-controllers-to-share-information/coronavirus-covid-19-notice-under-regulation-34-of-the-health-service-control-of-patient-information-regulations-2002-general--2). As of 1 July 2023, the Secretary of State has requested that NHS England continue to operate the Service under the COVID-19 Directions 2020. In some cases of data sharing, the common law duty of confidence is met using, for example, patient consent or support from the Health [Research Authority Confidentiality Advisory Group](https://www.hra.nhs.uk/about-us/committees-and-services/confidentiality-advisory-group/).

Taken together, these provide the legal bases to link patient datasets using the service. GP practices, which provide access to the primary care data, are required to share relevant health information to support the public health response to the pandemic and have been informed of how the service operates. This study was approved under the UK Statistics Authority ethics self-assessment review process (approval from the Health Research Authority was deemed as not required).

## Appendix

**Supplemental 1:**
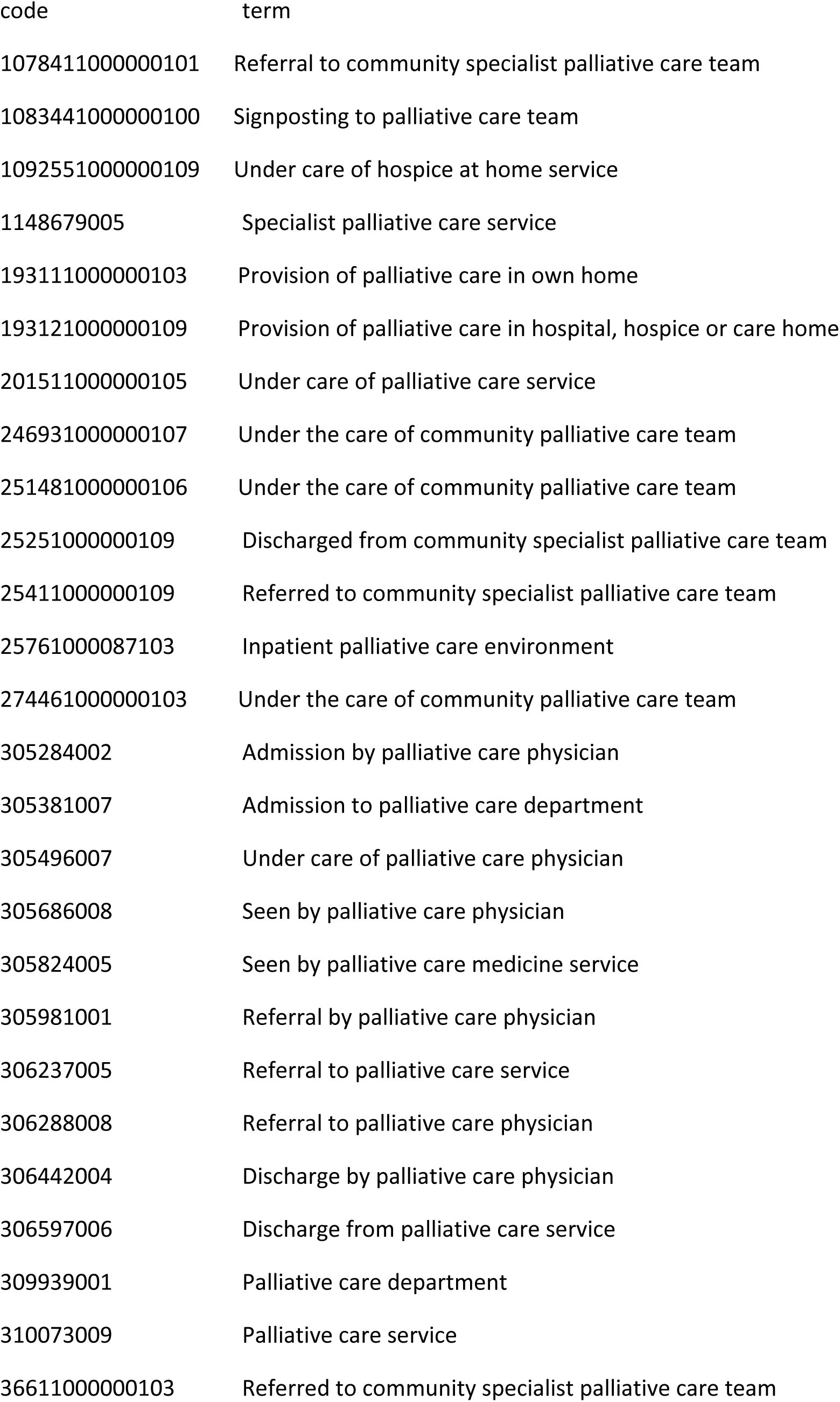
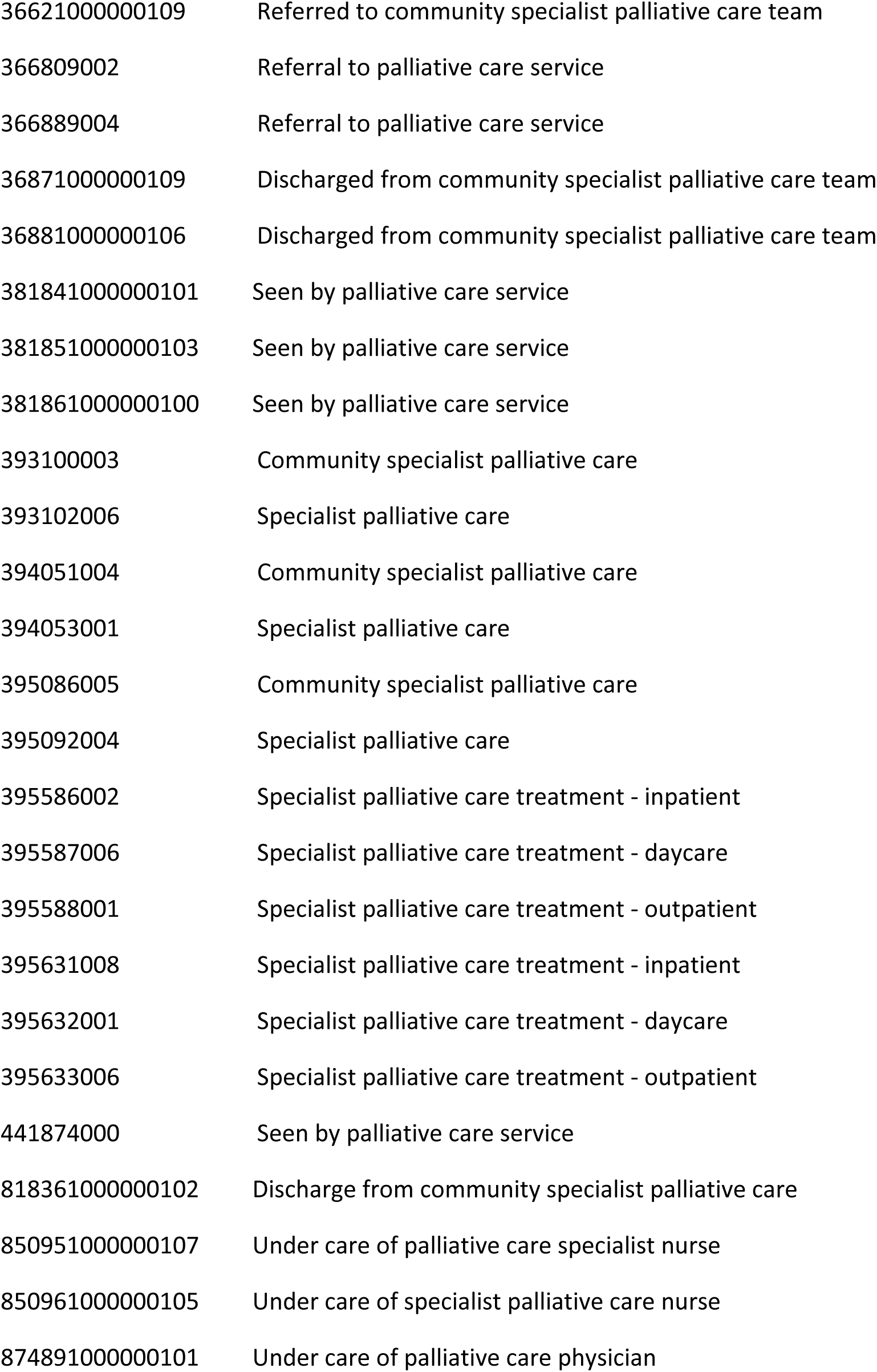
Codelist for specialist palliative care team contacts:

| code | term |
| --- | --- |
| 1078411000000101 | Referral to community specialist palliative care team |
| 1083441000000100 | Signposting to palliative care team |
| 1092551000000109 | Under care of hospice at home service |
| 1148679005 | Specialist palliative care service |
| 193111000000103 | Provision of palliative care in own home |
| 193121000000109 | Provision of palliative care in hospital, hospice or care home |
| 201511000000105 | Under care of palliative care service |
| 246931000000107 | Under the care of community palliative care team |
| 251481000000106 | Under the care of community palliative care team |
| 25251000000109 | Discharged from community specialist palliative care team |
| 25411000000109 | Referred to community specialist palliative care team |
| 25761000087103 | Inpatient palliative care environment |
| 274461000000103 | Under the care of community palliative care team |
| 305284002 | Admission by palliative care physician |
| 305381007 | Admission to palliative care department |
| 305496007 | Under care of palliative care physician |
| 305686008 | Seen by palliative care physician |
| 305824005 | Seen by palliative care medicine service |
| 305981001 | Referral by palliative care physician |
| 306237005 | Referral to palliative care service |
| 306288008 | Referral to palliative care physician |
| 306442004 | Discharge by palliative care physician |
| 306597006 | Discharge from palliative care service |
| 309939001 | Palliative care department |
| 310073009 | Palliative care service |
| 36611000000103 | Referred to community specialist palliative care team |

|  |  |  |
| --- | --- | --- |
| 1 |  |  |
| 2 |  |  |
| 3 | 36621000000109 | Referred to community specialist palliative care team |
| 4 |  |  |
| 5 | 366809002 | Referral to palliative care service |
| 6 |  |  |
| 7 | 366889004 | Referral to palliative care service |
| 8 |  |  |
| 9 | 36871000000109 | Discharged from community specialist palliative care team |
| 10 |  |  |
| 11 | 36881000000106 | Discharged from community specialist palliative care team |
| 12 |  |  |
| 13 | 381841000000101 | Seen by palliative care service |
| 14 |  |  |
| 15 | 381851000000103 | Seen by palliative care service |
| 16 |  |  |
| 17 | 381861000000100 | Seen by palliative care service |
| 18 |  |  |
| 19 | 393100003 | Community specialist palliative care |
| 20 |  |  |
| 21 | 393102006 | Specialist palliative care |
| 22 |  |  |
| 23 | 394051004 | Community specialist palliative care |
| 24 |  |  |
| 25 | 394053001 | Specialist palliative care |
| 26 |  |  |
| 27 | 395086005 | Community specialist palliative care |
| 28 |  |  |
| 29 | 395092004 | Specialist palliative care |
| 30 |  |  |
| 31 | 395586002 | Specialist palliative care treatment - inpatient |
| 32 |  |  |
| 33 | 395587006 | Specialist palliative care treatment - daycare |
| 34 |  |  |
| 35 | 395588001 | Specialist palliative care treatment - outpatient |
| 36 |  |  |
| 37 | 395631008 | Specialist palliative care treatment - inpatient |
| 38 |  |  |
| 39 | 395632001 | Specialist palliative care treatment - daycare |
| 40 |  |  |
| 41 | 395633006 | Specialist palliative care treatment - outpatient |
| 42 |  |  |
| 43 | 441874000 | Seen by palliative care service |
| 44 |  |  |
| 45 | 818361000000102 | Discharge from community specialist palliative care |
| 46 |  |  |
| 47 | 850951000000107 | Under care of palliative care specialist nurse |
| 48 |  |  |
| 49 | 850961000000105 | Under care of specialist palliative care nurse |
| 50 |  |  |
| 51 | 874891000000101 | Under care of palliative care physician |
| 52 |  |  |
| 53 |  |  |
| 54 |  |  |
| 55 |  |  |
| 56 |  |  |
| 57 |  |  |
| 58 |  |  |
| 59 |  |  |
| 60 |  |  |

**Supplemental 2:**
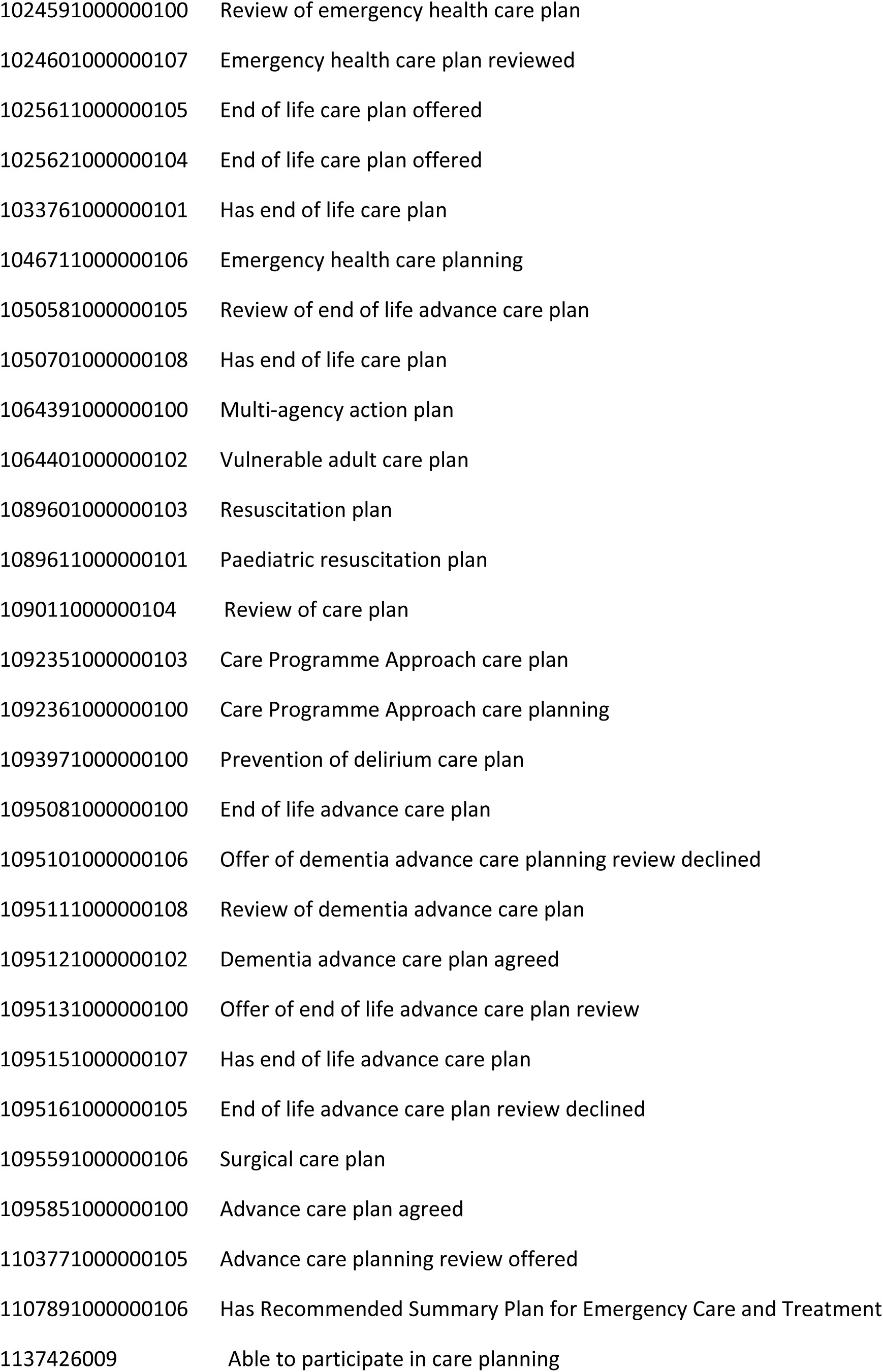
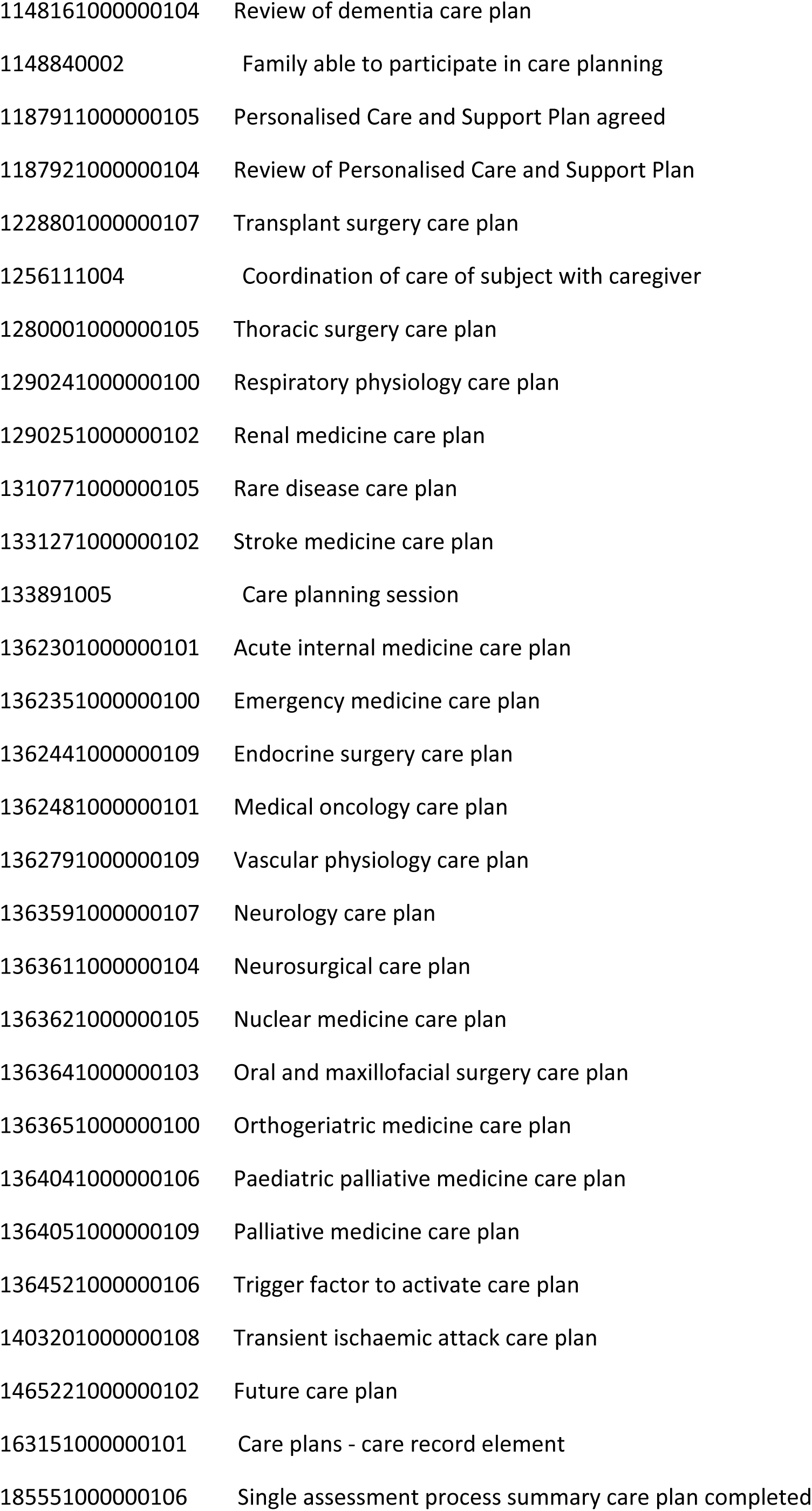
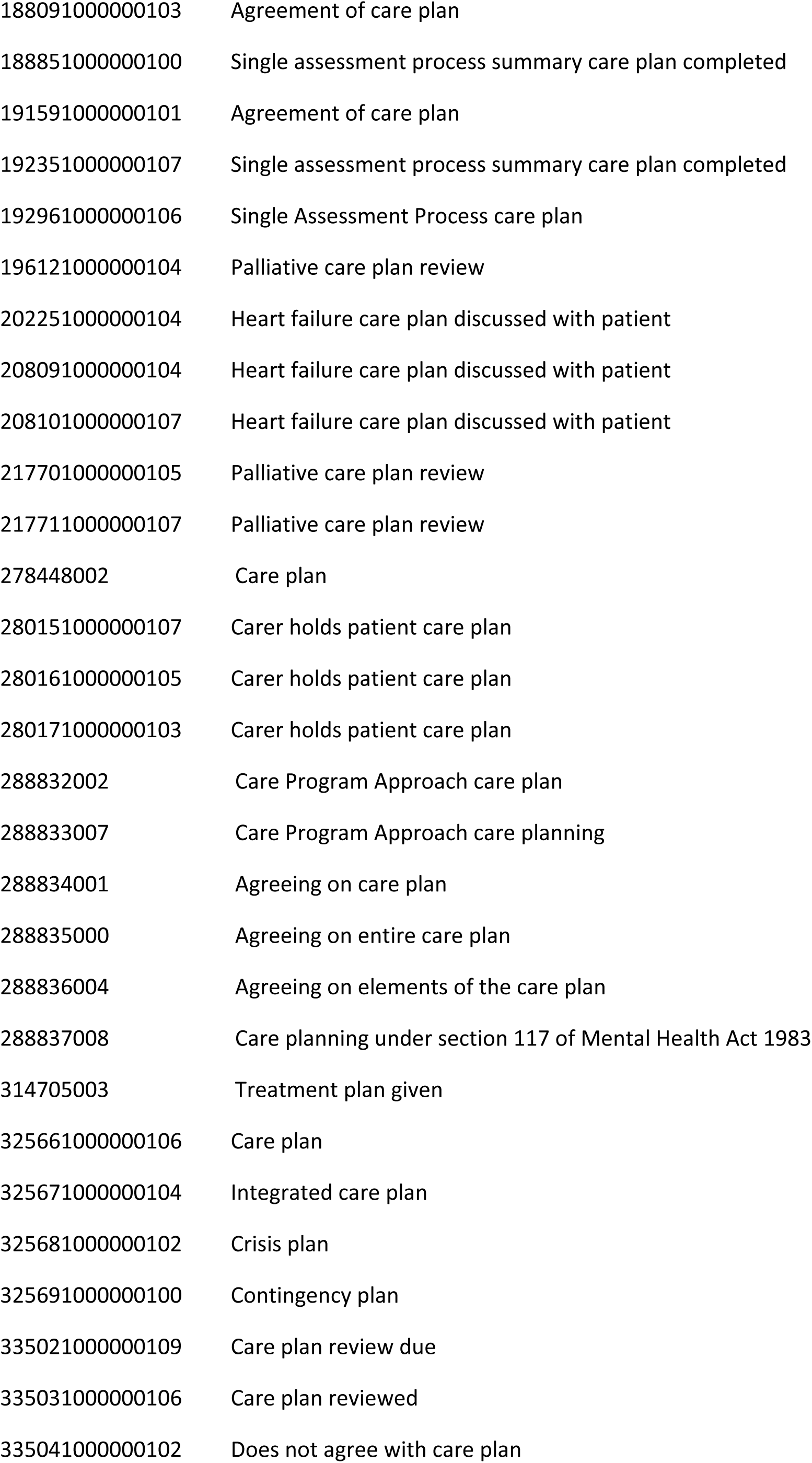
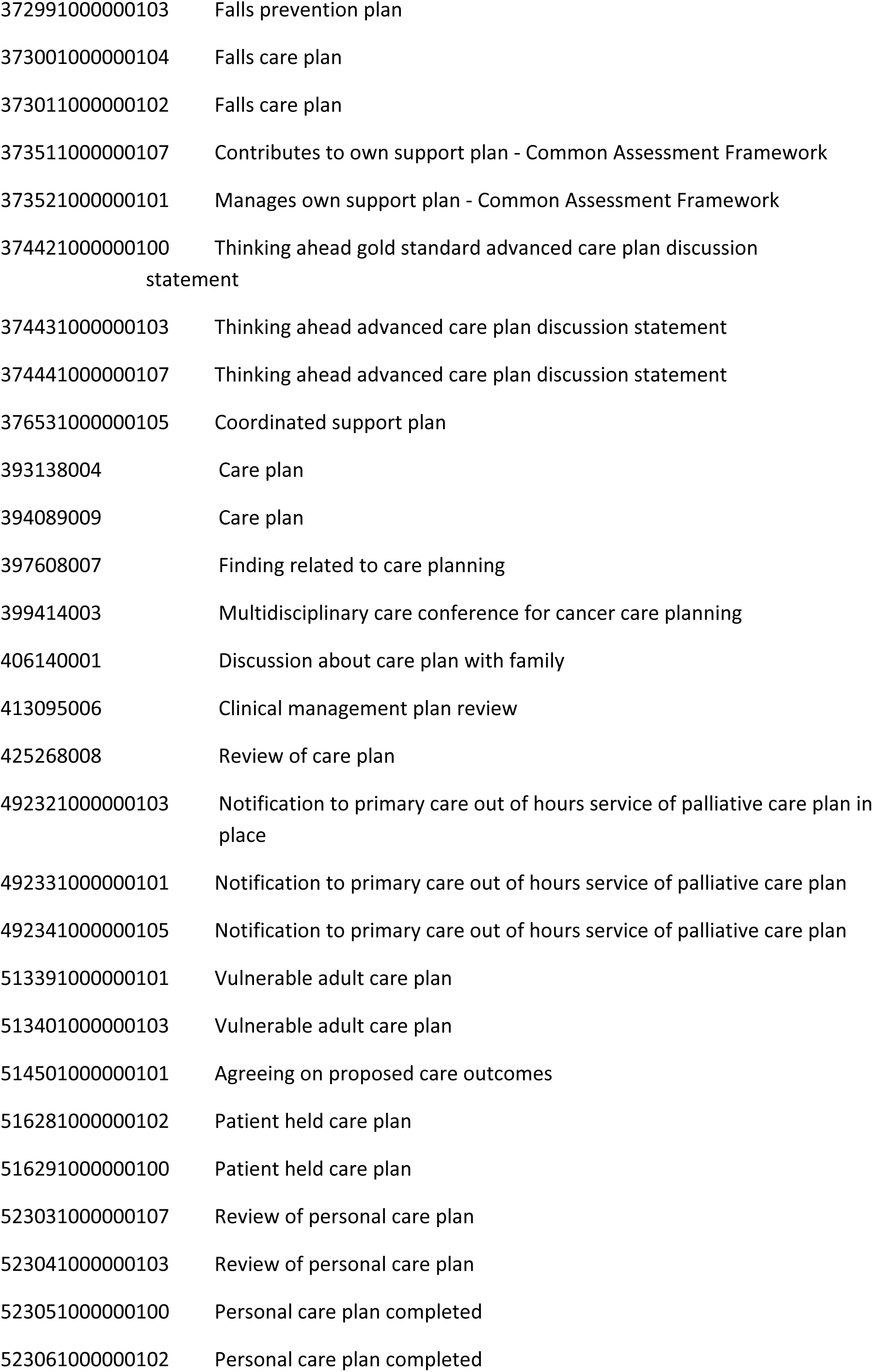
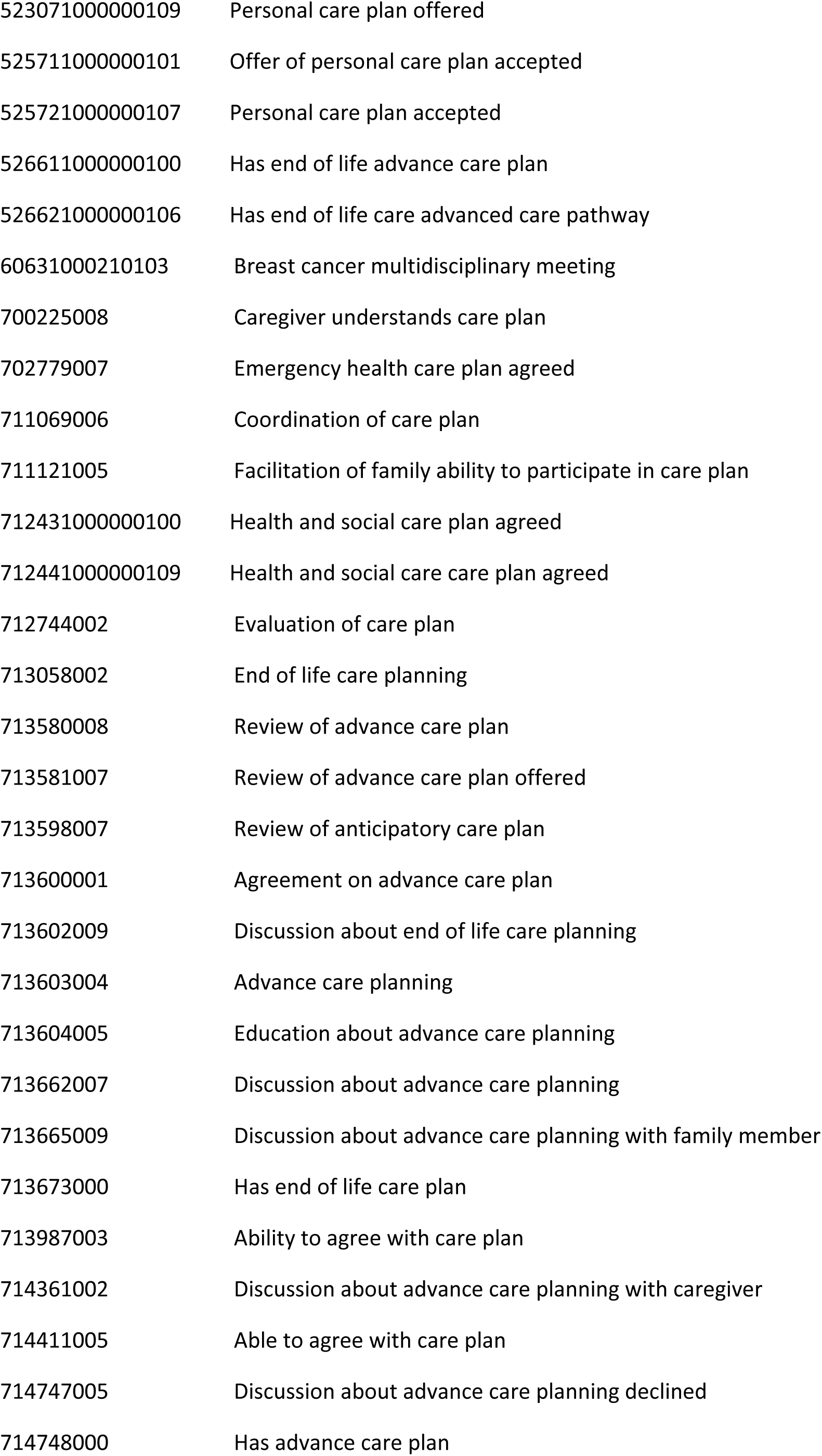
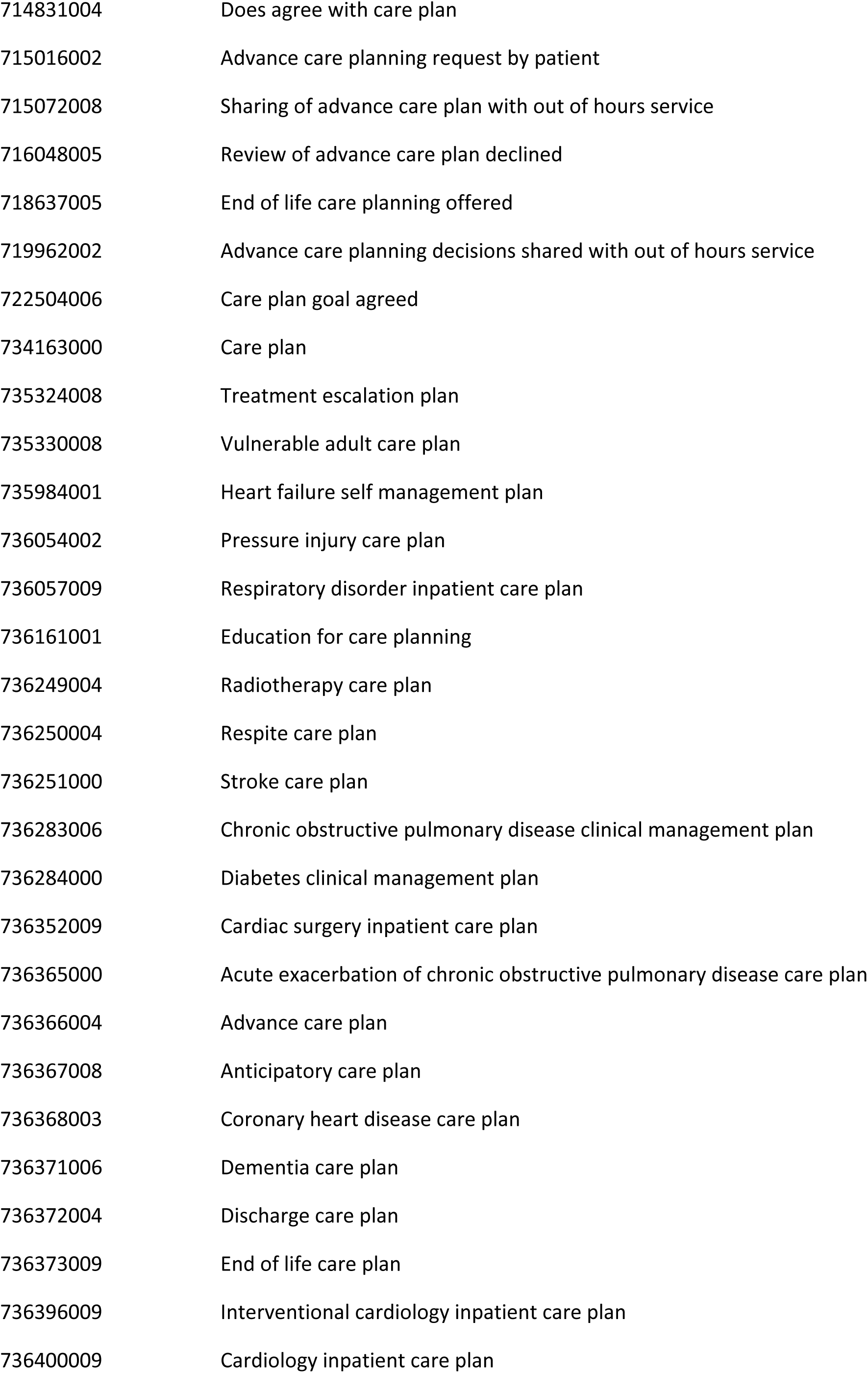
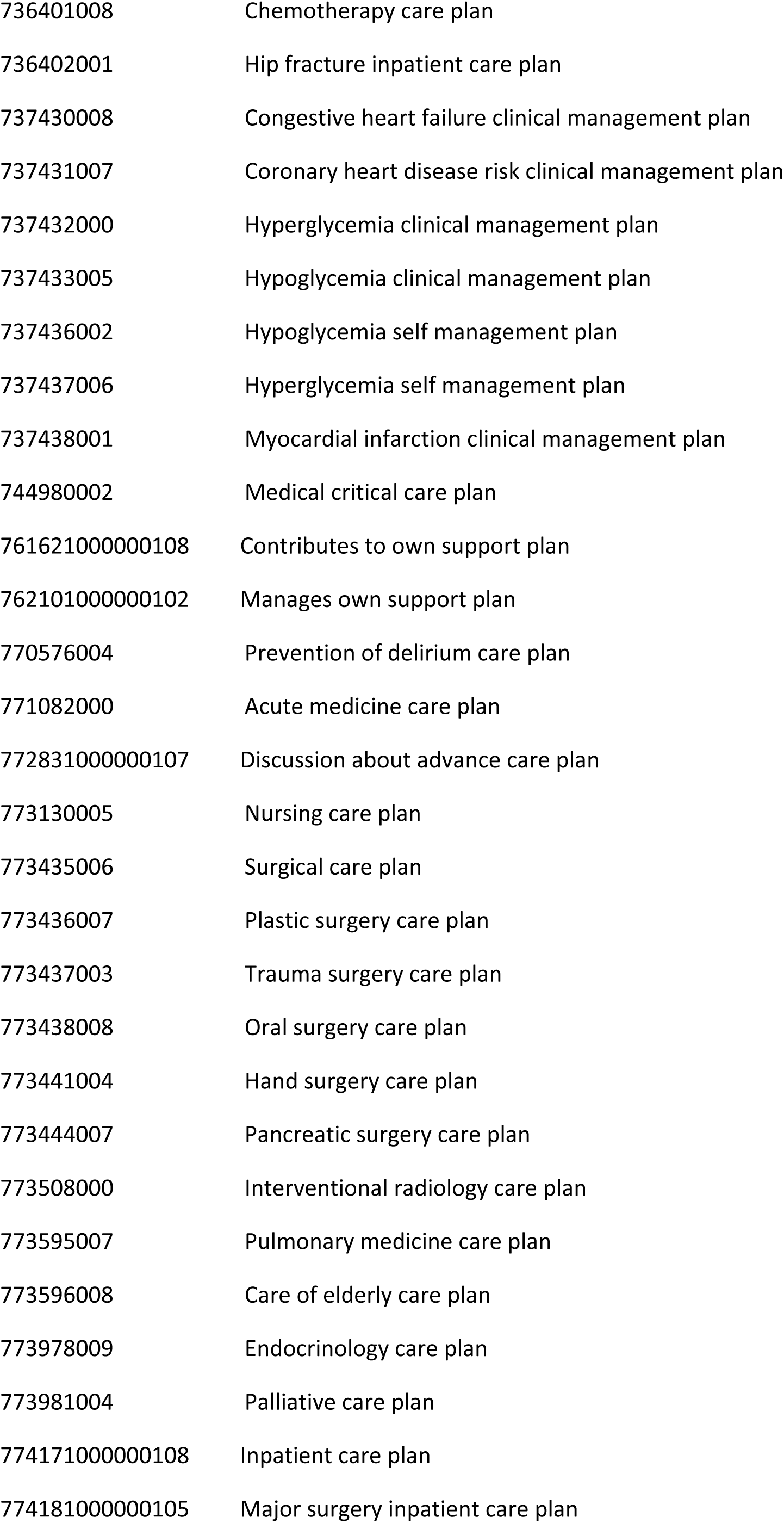
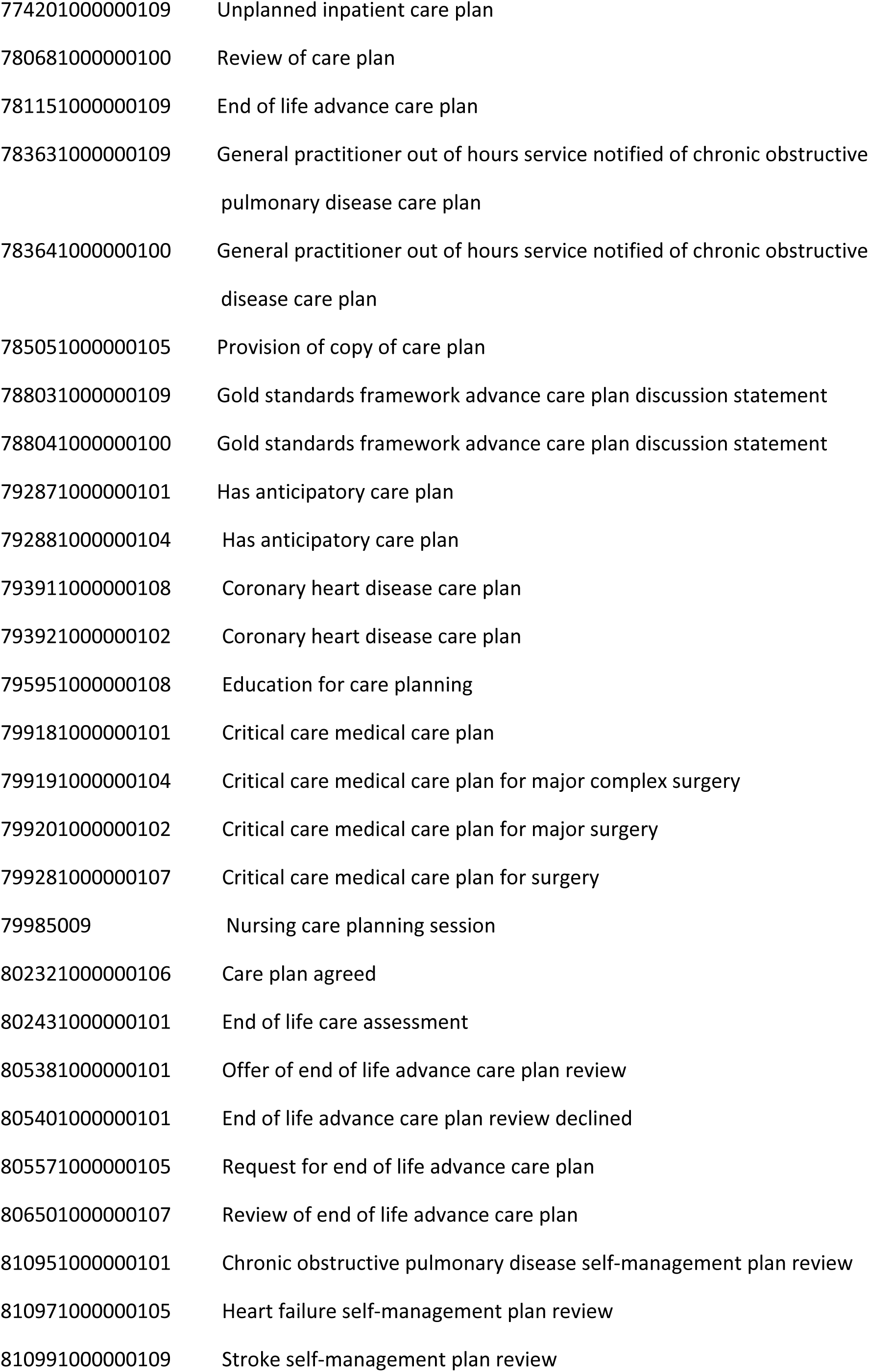
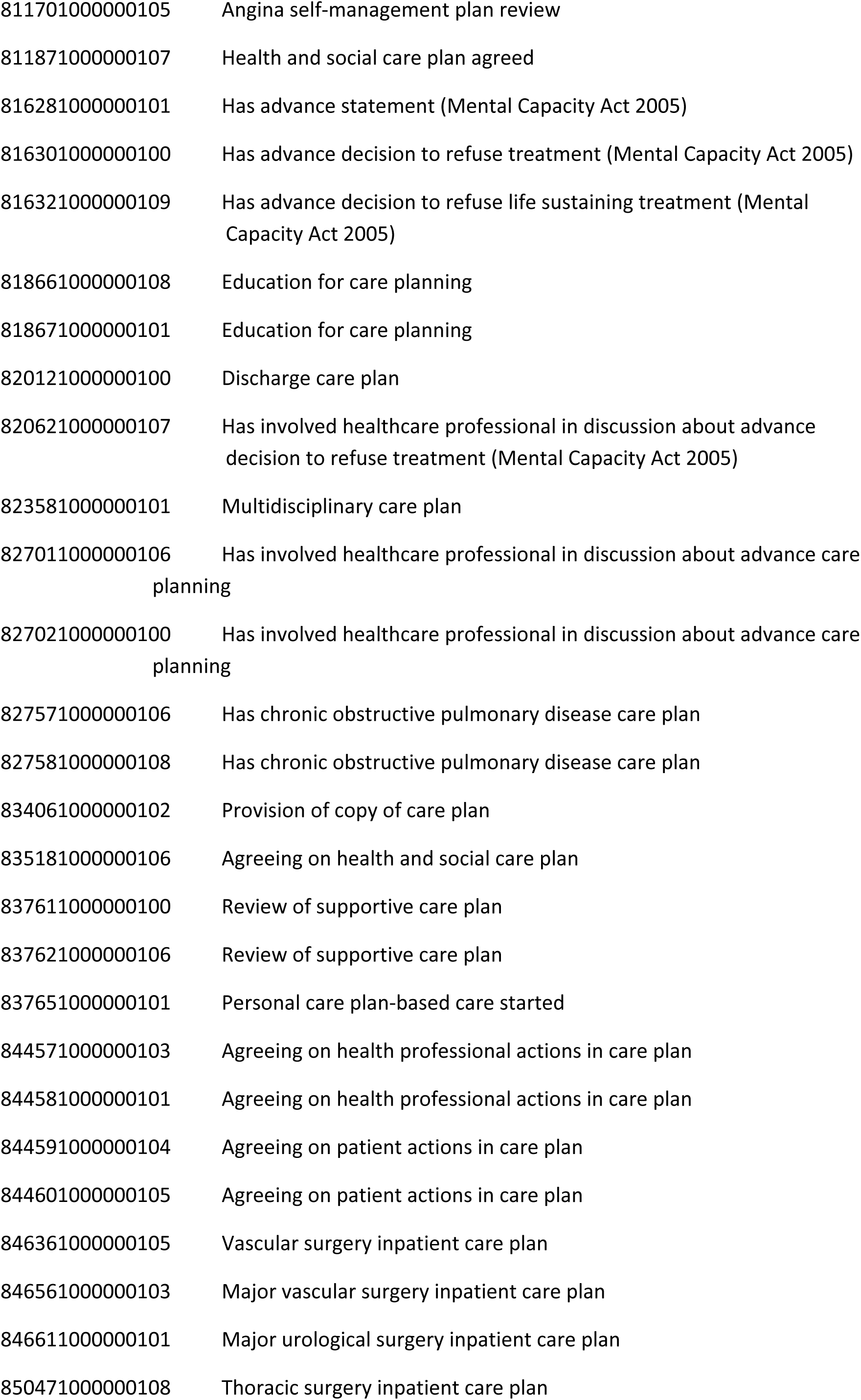
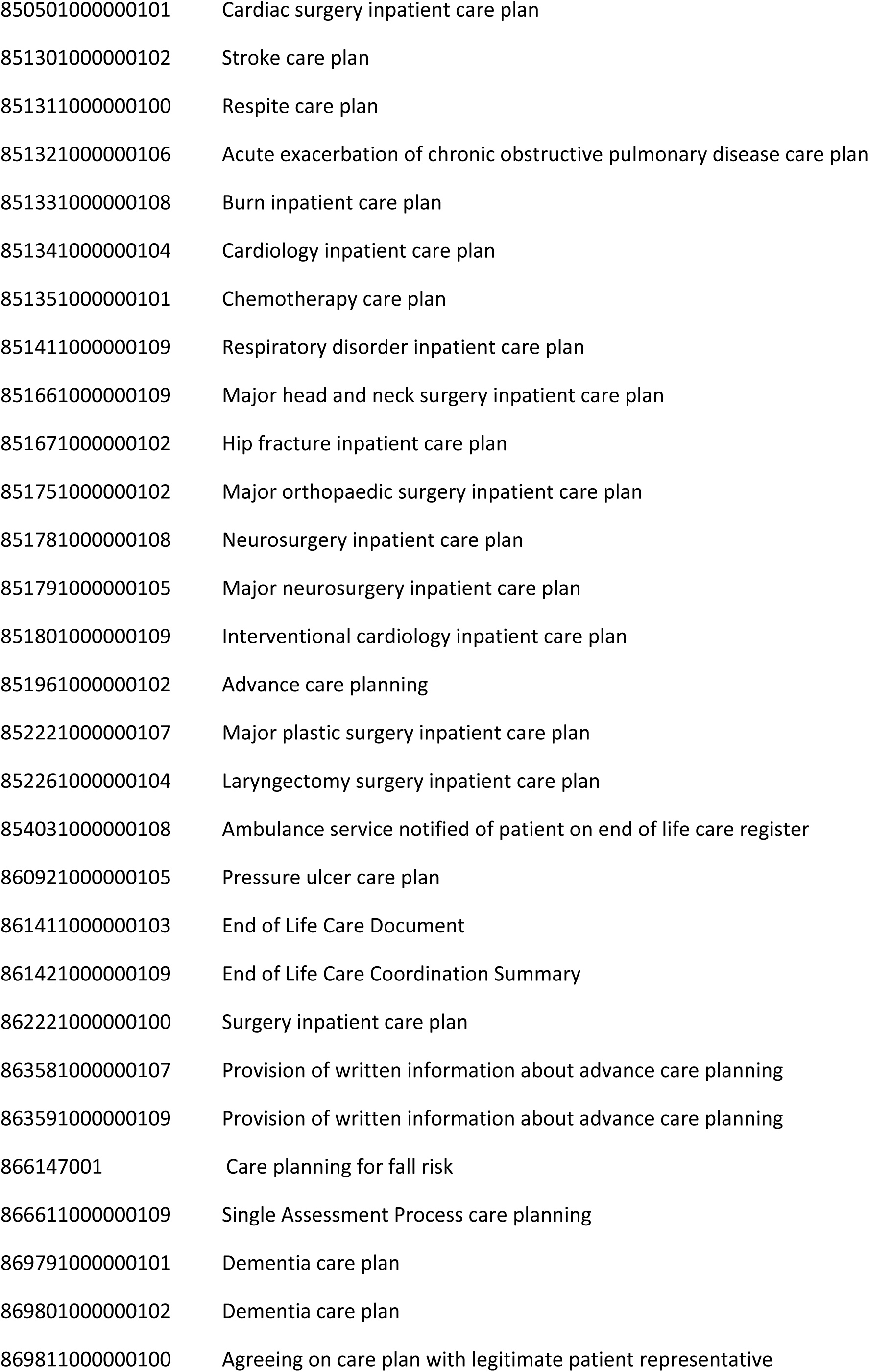
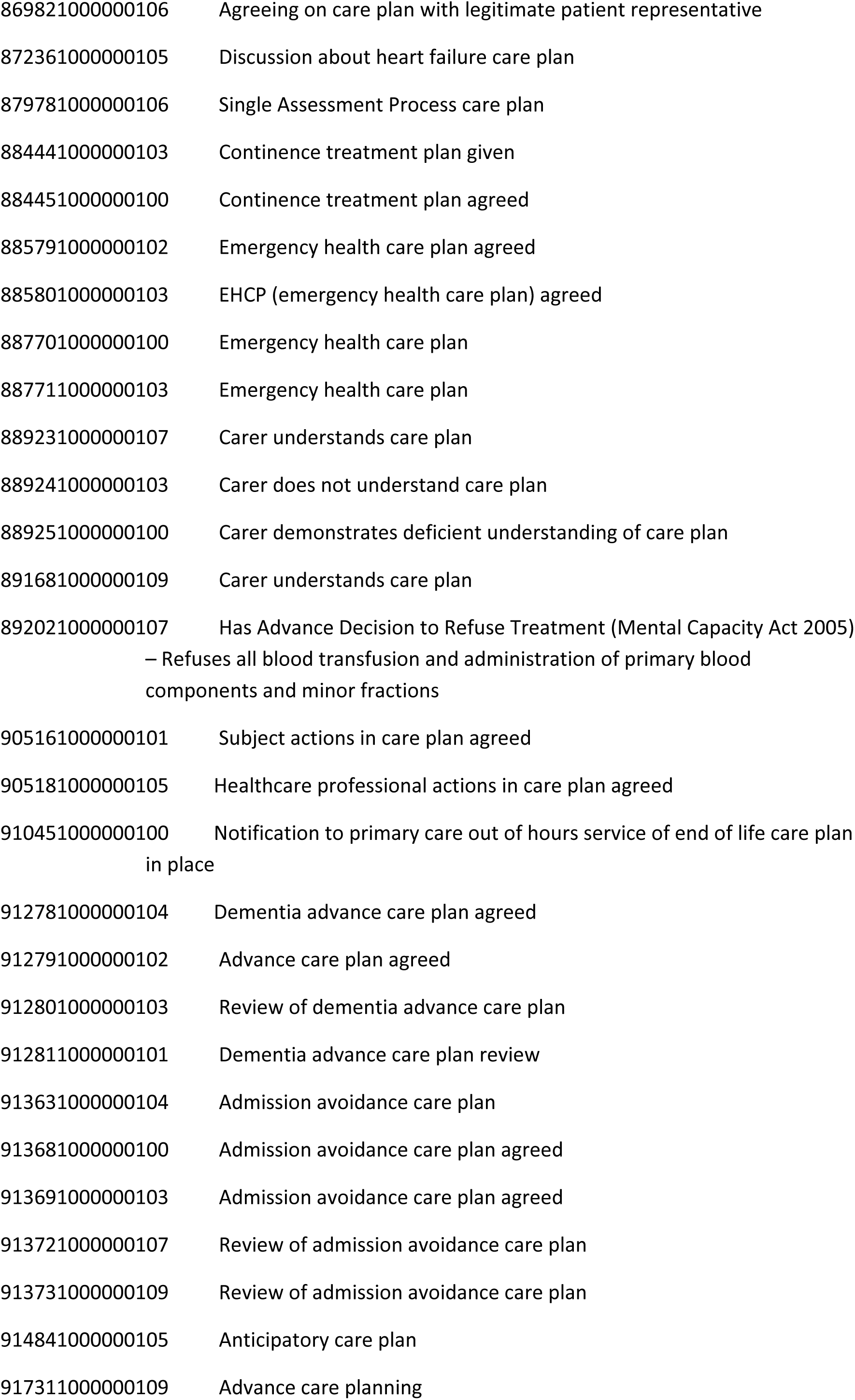
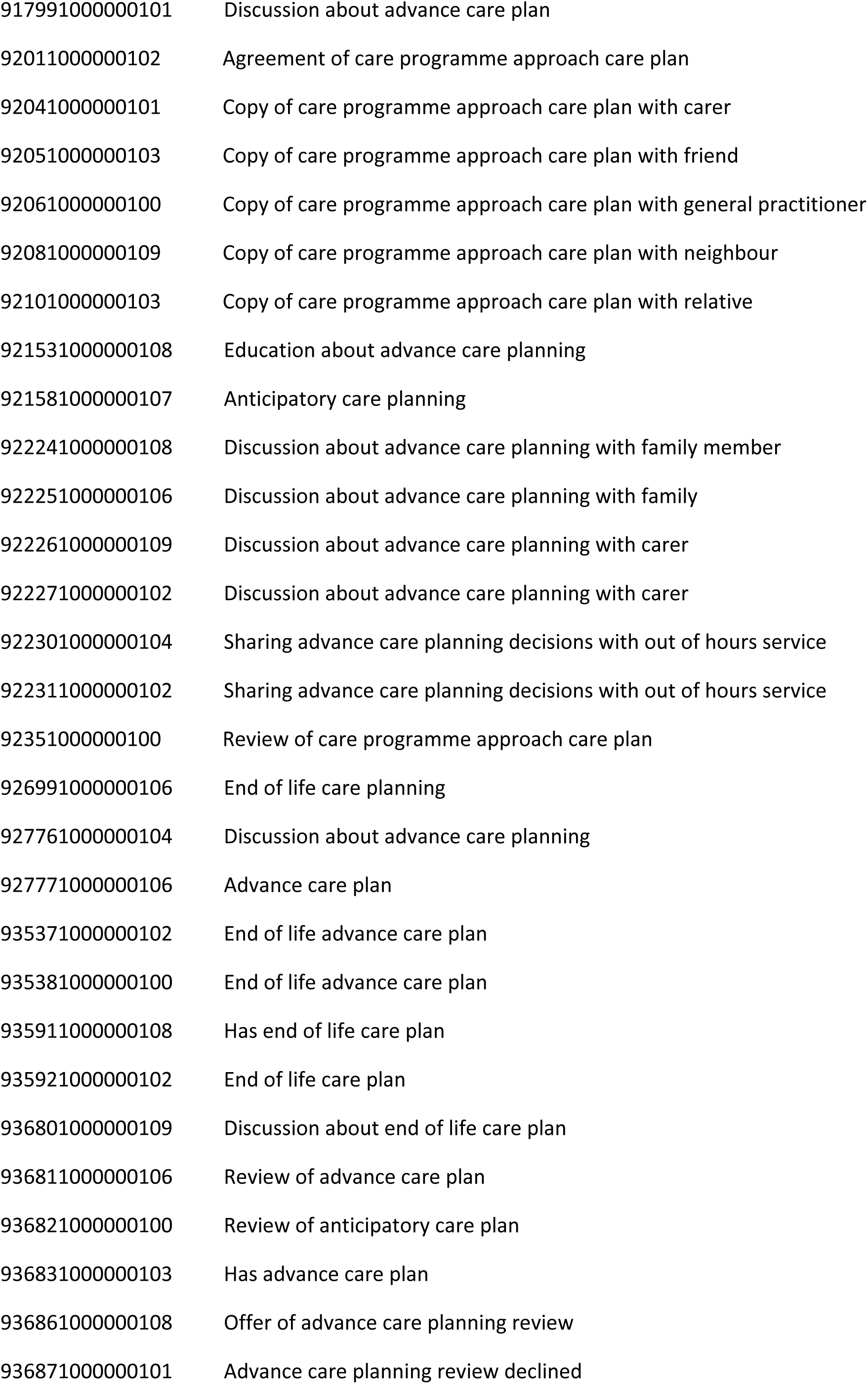
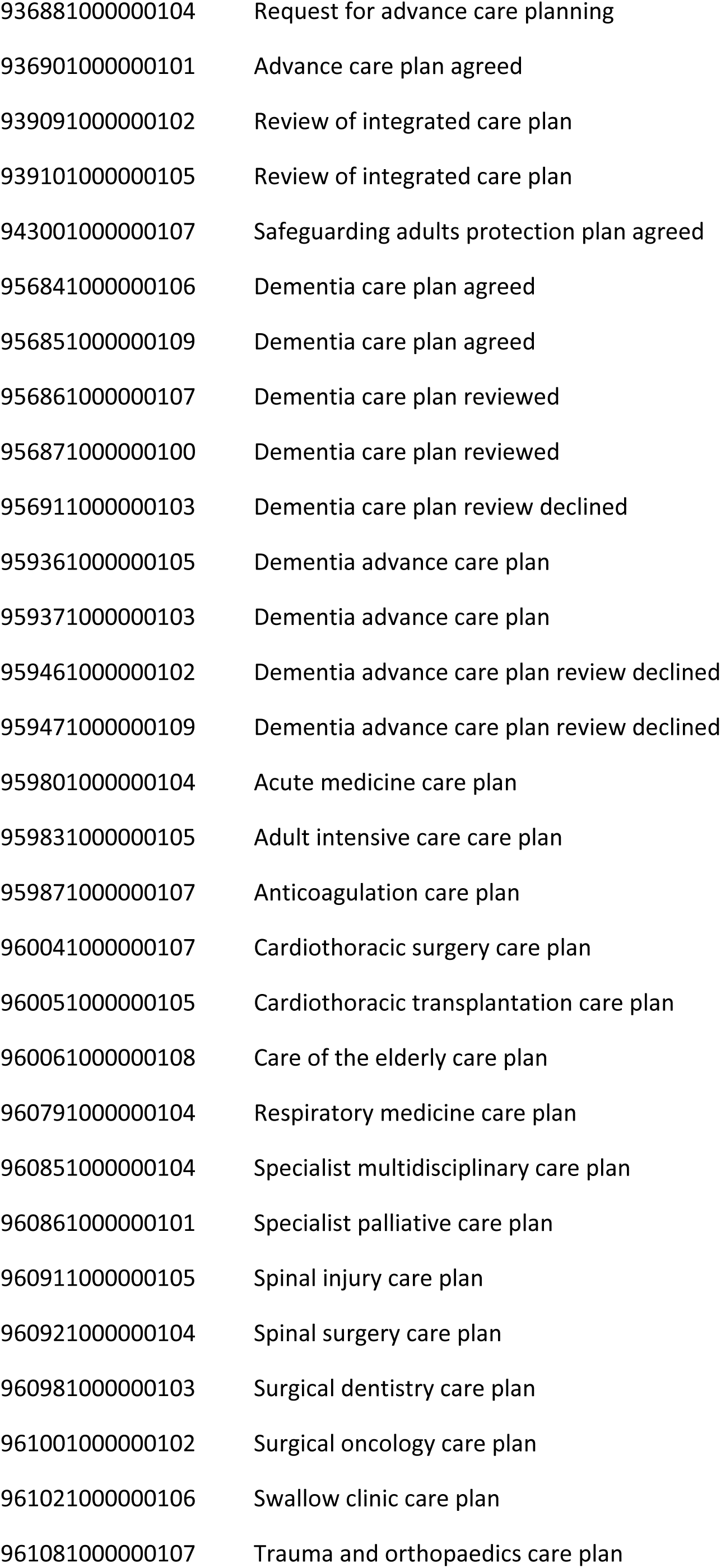
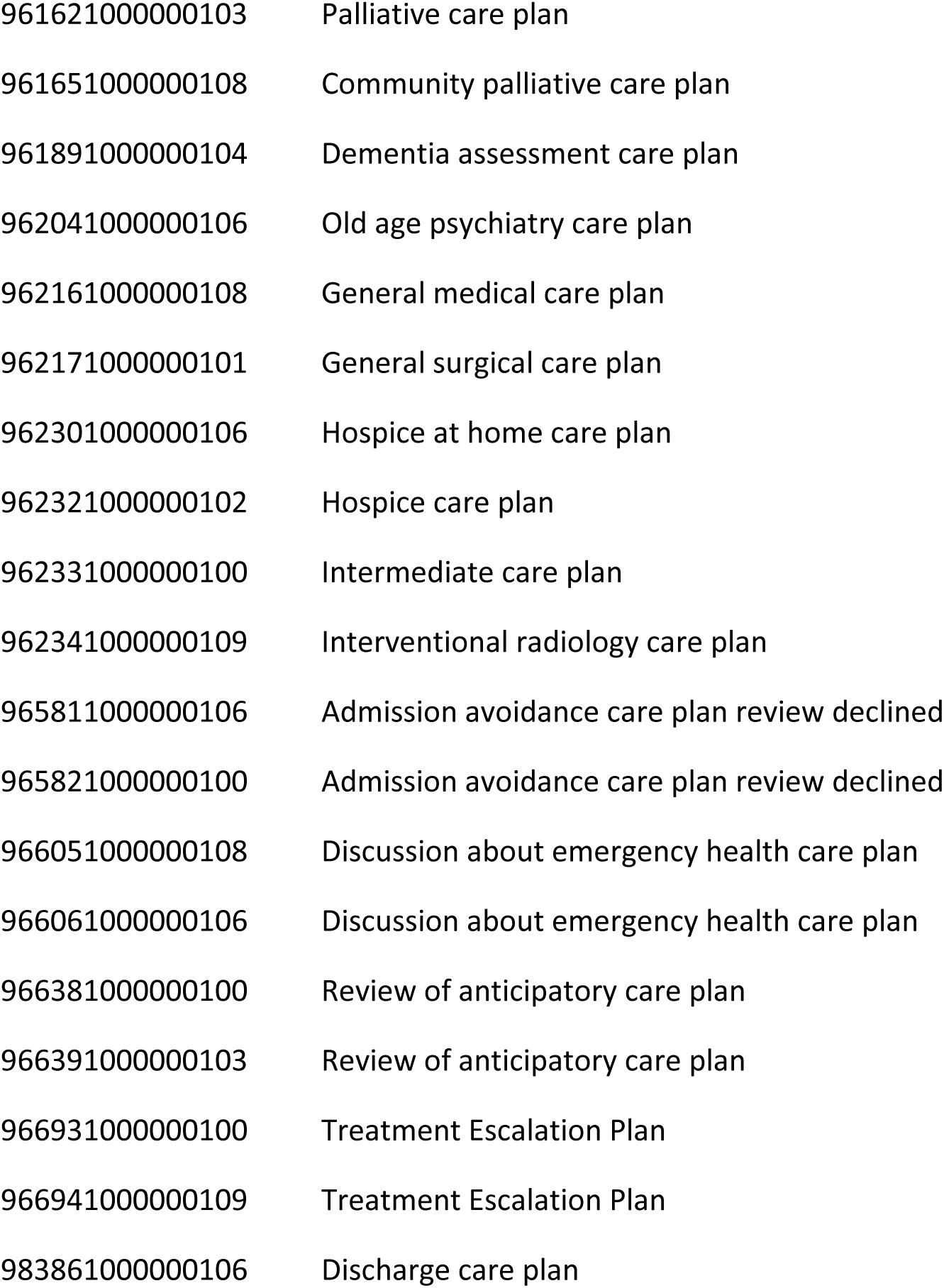
Codelist for advance care planning:

### Supplemental 3: A note on OpenSAFELY data

#### Data construction, linkage and verification

Access to the underlying identifiable and potentially re-identifiable pseudonymised electronic health record data is tightly governed by various legislative and regulatory frameworks and restricted by best practice. The data in the NHS England OpenSAFELY COVID-19 service is drawn from General Practice data across England where TPP is the data processor.

TPP developers initiate an automated process to create pseudonymised records in the core OpenSAFELY database, which are copies of key structured data tables in the identifiable records. These pseudonymised records are linked onto key external data resources that have also been pseudonymised via SHA-512 one-way hashing of NHS numbers using a shared salt. University of Oxford, Bennett Institute for Applied Data Science developers and PIs, who hold contracts with NHS England, have access to the OpenSAFELY pseudonymised data tables to develop the OpenSAFELY tools.

These tools in turn enable researchers with OpenSAFELY data access agreements to write and execute code for data management and data analysis without direct access to the underlying raw pseudonymised patient data, and to review the outputs of this code. All code for the full data management pipeline — from raw data to completed results for this analysis — and for the OpenSAFELY platform as a whole is available for review at github.com/OpenSAFELY.

## Notes

### Author Declarations

NHS England is the data controller of the NHS England OpenSAFELY COVID-19 Service; TPP is the data processor; all study authors using OpenSAFELY have the approval of NHS England. This implementation of OpenSAFELY is hosted within the TPP environment which is accredited to the ISO 27001 information security standard and is NHS IG Toolkit compliant. Patient data has been pseudonymised for analysis and linkage using industry standard cryptographic hashing techniques; all pseudonymised datasets transmitted for linkage onto OpenSAFELY are encrypted; access to the NHS England OpenSAFELY COVID-19 service is via a virtual private network (VPN) connection; the researchers hold contracts with NHS England and only access the platform to initiate database queries and statistical models; all database activity is logged; only aggregate statistical outputs leave the platform environment following best practice for anonymisation of results such as statistical disclosure control for low cell counts. The service adheres to the obligations of the UK General Data Protection Regulation (UK GDPR) and the Data Protection Act 2018. The service previously operated under notices initially issued in February 2020 by the Secretary of State under Regulation 3(4) of the Health Service (Control of Patient Information) Regulations 2002 (COPI Regulations), which required organisations to process confidential patient information for COVID-19 purposes; this set aside the requirement for patient consent. As of 1 July 2023, the Secretary of State has requested that NHS England continue to operate the Service under the COVID-19 Directions 2020. In some cases of data sharing, the common law duty of confidence is met using, for example, patient consent or support from the Health Research Authority Confidentiality Advisory Group. Taken together, these provide the legal bases to link patient datasets using the service. GP practices, which provide access to the primary care data, are required to share relevant health information to support the public health response to the pandemic and have been informed of how the service operates. This study was approved under the UK Statistics Authority ethics self-assessment review process (approval from the Health Research Authority was deemed as not required).

